# Association between the *LRP1B* and *APOE loci* and the development of Parkinson’s disease dementia

**DOI:** 10.1101/2022.05.23.22275465

**Authors:** Raquel Real, Alejandro Martinez-Carrasco, Regina H. Reynolds, Michael A. Lawton, Manuela M. X. Tan, Maryam Shoai, Jean-Christophe Corvol, Mina Ryten, Catherine Bresner, Leon Hubbard, Alexis Brice, Suzanne Lesage, Johann Faouzi, Alexis Elbaz, Fanny Artaud, Nigel Williams, Michele T. M. Hu, Yoav Ben-Shlomo, Donald G. Grosset, John Hardy, Huw R. Morris

## Abstract

Parkinson’s disease (PD) is one of the most common age-related neurodegenerative disorders. Although predominantly a motor disorder, cognitive impairment and dementia are important features of PD, particularly in the later stages of the disease. However, the rate of cognitive decline varies widely among PD patients, and the genetic basis for this heterogeneity is incompletely understood. Here, we have analysed 3,964 clinically diagnosed PD cases to explore the genetic factors associated with rate of progression to PD dementia. Genome-wide survival analysis identified the *APOE-*ε4 allele as a major risk factor for the conversion to PD dementia, as well as three new *loci*, including the ApoE and APP receptor *LRP1B.* Biomarker analysis also implicates the amyloid pathway in PD dementia, suggesting that amyloid-targeting therapy may have an important role in preventing PDD.

## Introduction

Parkinson’s disease (PD) is the second most common neurodegenerative disease, with an estimated worldwide prevalence of 100-200 cases per 100,000 individuals^1^. Although PD is mainly viewed as a motor disorder, the development of dementia in PD is an important determinant of morbidity, mortality and need for social support.^2^ The clinico-pathological phenotype of Parkinson’s disease dementia (PDD) can be indistinguishable from dementia with Lewy bodies (DLB), although in PDD motor symptoms must by definition precede the development of dementia by at least one year^3^. Neuropsychiatric manifestations of PDD include cognitive fluctuation with visual misperception, hallucinations, and delusions together with deficits in attention, executive and visuo-spatial function. Cholinergic denervation seems to be important in PDD and cholinesterase inhibitors can improve PDD symptoms^4^, but there is no treatment for the underlying disease pathology. Identifying the causal mechanisms will be an important step in defining new treatments.

Age is the single most important risk factor for PDD. It is estimated that by the age of 90, 80-90% of individuals with PD will have developed dementia.^4^ Other clinical predictors of progression to dementia include disease severity and longer disease duration^5–7^. However, the rates of progression to PDD vary substantially among individuals, which has important implications for prognosis and quality of life. Several genetic factors have been reported to increase the risk or rate of progression to PDD. The most widely reported genetic risk factor associated with increased risk of conversion to PDD is the *APOE* ε4 allele^8–10^. Single rare variants in the *GBA* gene increase the risk of developing PDD, and the risk may relate to the pathogenicity of the variant^11–13^. Several studies have also reported that the *MAPT* H1 haplotype is associated with dementia^9, 14, 15^, although this has not been universally replicated^10^. Genome-wide association studies in neurodegenerative disease have largely defined case-control risk factors for disease susceptibility, but the increasing availability of high-quality longitudinal clinical datasets enables a systematic search for disease modifying factors. Here, we use a genome-wide survival analysis approach to identify new genetic factors that contribute to the progression to PDD.

## Results

### Cohort characterisation

Following data cleaning (Supplementary Fig. 1), a total of 3,964 individuals diagnosed with PD were available for analysis from the Tracking Parkinson’s (TPD)^16^, Oxford Parkinson’s Disease Centre Discovery Cohort (OPDC)^17^, Accelerating Medicines Partnership: Parkinson’s Disease (AMP-PD)^18^, and Drug Interaction With Genes in Parkinson’s Disease (DIGPD). Demographic characteristics of each patient cohort are shown in Supplementary Table 1. Participants in DIGPD and AMP-PD cohorts were significantly younger at PD onset (Kruskal-Wallis chi-squared value = 198.99, df = 3, post hoc Dunn’s multiple comparison test, TPD vs DIGPD: *P* = 2.08x10^-19^; TDP vs AMP-PD: *P* = 1.09x10^-27^; OPDC vs DIGPD, *P* = 2.35x10^-17^; OPDC vs AMP-PD: *P* = 6.53x10^-21^) and at study baseline (Kruskal-Wallis chi-squared value = 159.86, df = 3, post hoc Dunn’s multiple comparison test, TPD vs DIGPD: *P* = 3.07x10^-18^; TDP vs AMP-PD: *P* = 1.11x10^-21^; OPDC vs DIGPD: *P* = 4.98x10^-15^; OPDC vs AMP-PD: *P* =1.10x10^-14^), which is probably reflected in the significantly reduced event rates in these two cohorts (χ^2^ test with Yates correction, TPD *vs* AMP-PD: χ^2^ = 17.6, df = 1, *P* = 2.758x10^-05^; OPDC *vs* AMP-PD: χ^2^ = 61.8, df = 1, *P* = 3.883x10^-15^; OPDC *vs* DIGPD: χ^2^ = 14.7, df = 1, *P* = 1.287x10^-04^). Adjusted MoCA or MMSE scores over time in PD cases who never developed dementia during the study follow-up remained constant over time, while they were consistently lower and showed greater decline in individuals who went on to develop PDD during study follow-up (Supplementary Fig. 2).

### Identification of genetic determinants of PDD

In our genome wide survival meta-analysis assessing the role of 6,107,418 single nucleotide polymorphisms (SNPs) in the development of PDD, we identified four genome-wide significant genetic *loci* (Fig. 1 and Table 1; regional association plots in Supplementary Fig. 3). The most significant SNP was the ε4 allele-tagging SNP rs429358 in *APOE* (Hazard Ratio (HR) = 2.42; 95% Confidence Interval (CI) = 1.95, 3.01; *P* = 1.2 x10^-15^). *APOE* is the most important genetic risk factor for the development of Alzheimer’s disease (AD) and has also been shown in multiple studies to contribute to cognitive decline and dementia in PD^8–10, 19^. Conditional analysis on the lead SNP at the *APOE locus* did not reveal any other independent SNPs contributing to the signal at this location (Supplementary Fig. 4a-b and Supplementary Table 2).

**Fig. 1.**
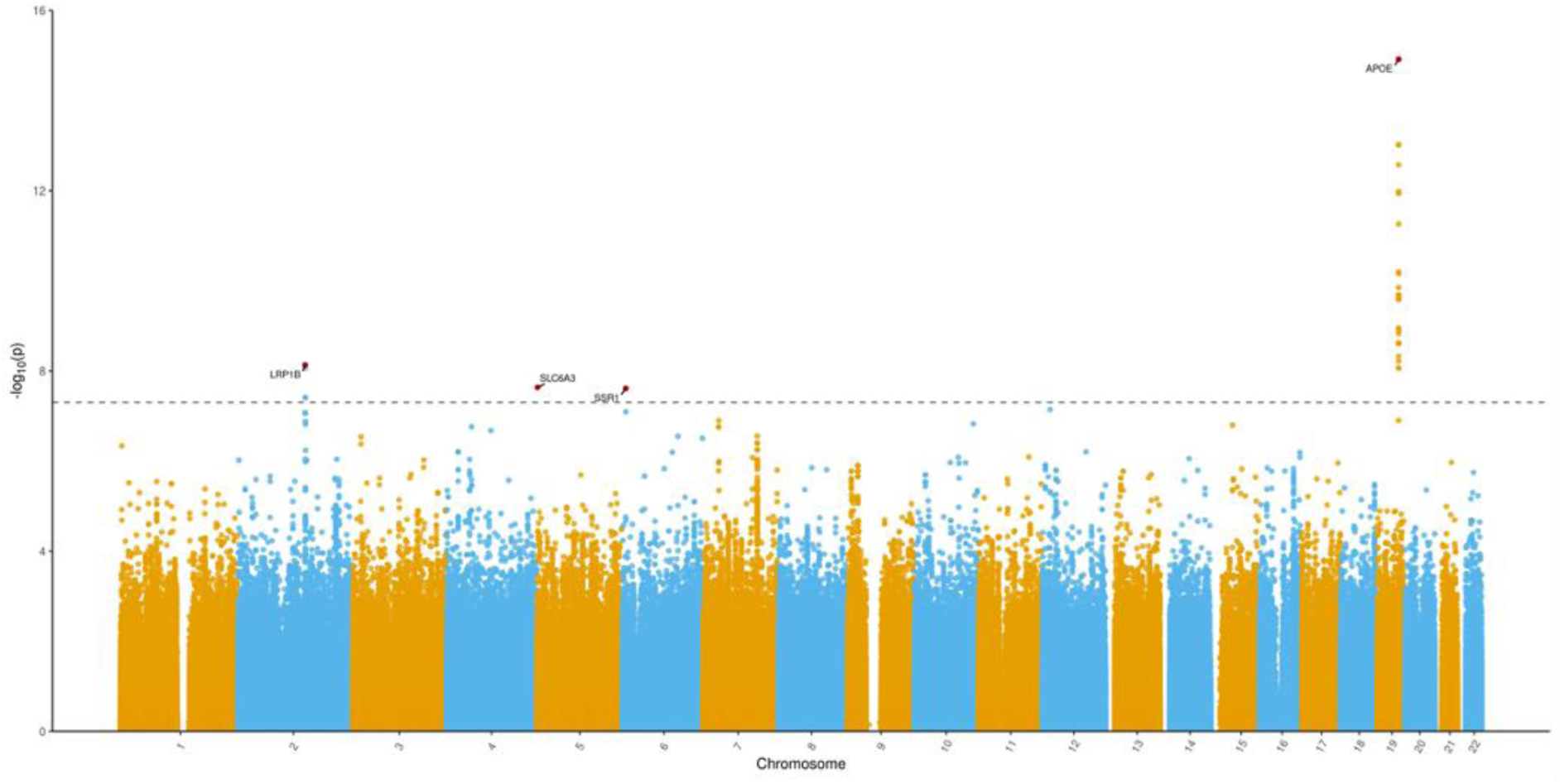
Manhattan plot representing the results of the GWSS meta-analysis. The GWSS was conducted using a Cox proportional hazards model in each cohort separately, and results were meta-analysed (PDD: *n* = 272; PD: *n* = 3,692). Red dots indicate the variant with the lowest *P*-value at each genome-wide significant genetic *locus* and the gene closest to the top variant is indicated. Genome-wide significance was set at 5x10^-8^ and is indicated by the dashed line.

**Table 1.** Top independent SNPs from GWSS meta-analysis

| CHR | BP | SNP ID | Effect allele | Nearest gene | Effect allele frequency |  |  | HR | 95% CI | P-value |
| --- | --- | --- | --- | --- | --- | --- | --- | --- | --- | --- |
|  |  |  |  |  | PD | PDD | NFE |  |  |  |
| 19 | 45411941 | rs429358 | C | *APOE* | 0.1813 | 0.2224 | 0.1486 | 2.42 | 1.95-3.01 | 1.21x10<sup>-15</sup> |
| 2 | 142000271 | rs80306347 | C | *LRP1B* | 0.0214 | 0.0515 | 0.0277 | 3.37 | 2.23-5.09 | 7.39x10<sup>-09</sup> |
| 5 | 1415068 | rs28363070 | A | *SLC6A3* | 0.0123 | 0.0257 | 0.0131 | 4.42 | 2.62-7.45 | 2.32x10<sup>-08</sup> |
| 6 | 7320911 | rs60707664 | A | *SSR1* | 0.0508 | 0.0993 | 0.0536 | 2.31 | 1.72-3.11 | 2.44x10<sup>-08</sup> |

CHR, chromosome; BP, base pair position in hg19; HR, hazard ratio; 95% CI, 95% confidence interval, NFE, non-Finnish European from gnomAD (https://gnomad.broadinstitute.org/). Genome-wide significance level set at 5x10<sup>-08</sup>.

The second genome-wide significant genetic *locus* was on chromosome 2, in a region spanning ∼633kb, as defined by SNPs in LD with the lead SNP (r^2^ > 0.6). The lead SNP at this locus was rs80306347 (HR = 3.37; 95% CI = 2.23-5.09; *P* = 7.39x10^-09^). This is an intronic variant located in intron 5 of the *LRP1B* gene (ENSG00000168702). A second genome-wide significant SNP nominated as independent by FUMA (given the r^2^ < 0.6)^20^ was present at this *locus* (HR = 3.48; 95% CI = 2.28-5.32; *P* = 8.49x10^-09^). rs116725215 is also an intronic variant located in intron 2 of *LRP1B*. This gene encodes the low-density lipoprotein (LDL) receptor-related protein 1B, a member of the LDL receptor superfamily. LRP1B is a receptor for ApoE-carrying lipoproteins and is highly expressed in the adult human brain (Supplementary Fig. 5a,d)^21^. In addition, *LRP1B* was found to be significantly upregulated in excitatory neurons of the anterior cingulate cortex of PDD compared to PD and control brain samples (Supplementary Fig. 6)^22^. Similar to other LDL receptors, it is involved in the intracellular processing of the amyloid precursor protein (APP)^23^. Therefore, *LRP1B* constitutes a promising candidate for regulating the development of dementia in PD. Conditional analysis did not confirm independence of variants rs80306347 and rs116725215, as conditioning on either SNP abolished genome-wide significance at the *LRP1B locus* (Supplementary Fig. 4c-d and Supplementary Table 3). rs28363070 is an intronic variant in the *SLC6A3* gene (ENSG00000142319) in chromosome 5 and was associated with progression to dementia with genome-wide significance (HR = 4.42; 95% CI = 2.62-7.45; *P* = 2.32x10^-08^). *SLC6A3* encodes the dopamine transporter (DAT) highly expressed in the brain, specifically in nigro-striatal neurons (Supplementary Fig. 5b). It controls dopamine reuptake at the presynaptic terminals of dopaminergic neurons, thereby regulating dopamine availability in the striatum. Given its prominent role in the metabolism of dopamine, there has been long-standing interest in this gene in relation to the pathophysiology of PD. For example, while genome-wide association studies have not found an association between *SLC6A3* variants and the risk of PD, the 9-repeat (9R) allele of the 40-base pair variable number of tandem repeats (VNTR) polymorphism in the 3’-UTR region (rs28363170) has been shown to increase dopamine binding in the striatum, which could in theory confer increased vulnerability to neurotoxicity by oxidation due to enhanced dopamine neuronal uptake^24, 25^. Interestingly, PD cases carrying the 9R allele have been shown to perform worse on cognitive tasks linked to working memory and executive functions, and decreased activation of cortico-striatal loops while planning a set-shift task^26^. The effect of the *SLC6A3* in this study is heterogeneous in that only one of the cohorts contributed to this signal (Supplementary Fig. 7d), and thus the meta-analysis does not support a definite association of variation at *SLC6A3* with the development of dementia in PD.

Finally, variant rs60707664 on chromosome 6 was associated with progression to dementia (HR = 2.31; 95% CI = 1.72-3.11; *P* = 2.44x10^-08^). This variant is intergenic and *SSR1* (ENSG00000124783) is the nearest gene. *SSR1* is a ubiquitously expressed gene (Supplementary Fig. 5c) that encodes the translocon-associated protein subunit alpha (TRAPA), one of the four subunits of the endoplasmic reticulum (ER) membrane receptor complex TRAP that binds calcium to the ER membrane and is associated with regulation of protein translocation across the ER membrane. Interestingly, ER dysfunction has been shown to contribute to the pathogenesis of many neurodegenerative diseases including PD^27, 28^.

Forest plots of the GWSS meta-analysis (Supplementary Fig. 7a-d) show that effect size and direction are consistent for the *APOE* and *SSR1 loci* across all cohorts. In the *LRP1B locus*, a strong association with progression to PDD was observed in the TPD and OPDC cohorts, but not in AMP-PD (note that due to reduced number of events in individuals in the DIGPD cohort, infinite estimates were generated by the Cox proportional hazards analysis of this cohort). Several factors could be contributing to these differential observations between cohorts, namely the reduced event rate in the AMP-PD cohort (3.61%) compared to TPD and OPDC (7.3% and 12.7%, respectively). This could in turn be related to the younger age at baseline and shorter follow-up times in the AMP-PD cohorts, since increasing age is the most significant clinical risk factor for the development of dementia in PD (Supplementary Table 1). To evaluate the effect of the different event rates on the power to detect a genome-wide significant effect on dementia-free survival, we modelled statistical power for a hypothetical SNP with minor allele frequency 0.02 and an hazard ratio of 2, assuming a median time to the event of 4.5 years, under an additive genetic risk model (Supplementary Fig. 8). The sample size required to detect such a SNP with genome-wide significance at 80% statistical power is approximately 50% and 70% larger at the event rate observed in the AMP-PD cohort than at the event rates observed in the TPD and OPDC cohorts, respectively, thus demonstrating how a low event rate can hinder the ability of the survival analysis to detect significant effects of variants with rare minor allele frequencies. We also explored the possibility that heterogeneity in the AMP-PD dataset could be contributing to an apparent discrepancy of results across cohorts. AMP-PD is a clinically and genetically harmonised cohort, but each individual study has distinct design and inclusion criteria. We therefore analysed the *LRP1B locus* in each of the studies independently, and subsequently meta-analysed the results. Given the study design and reduced sample size of the independent studies, not all of them contributed to the meta-analysis. For example, the BioFIND samples did not include PDD cases, thus being excluded from the survival analysis. In the case of SURE-PD3, the low event rate (1.3%) combined with the rare frequency of the LRP1B allele meant that no PDD cases carried the allele of interest, therefore generating infinite estimates in the Cox proportional hazards analysis. Despite the decrease in power that comes with analysing smaller cohorts, the meta-analysis again showed that the LRP1B variant rs80306347 was associated with progression to dementia with genome-wide significance (HR = 3.88; 95% CI = 2.55-5.87; *P* = 1.609x10^-10^) and the forest plot (Supplementary Fig. 7e) demonstrates that effect size and direction are consistent across all studies that have contributed to the meta-analysis (i.e., from which valid estimates were generated).

### Colocalization analysis

We did not identify proxy coding variants in high linkage disequilibrium (LD) with the lead variants in *LRP1B*, *SLC6A3* and *SSR1*. To determine if any of the GWSS genome-wide significant signals are involved in the regulation of gene expression, we performed colocalization analysis using expression quantitative trait *loci* (eQTLs) from eQTLGen^29^ and PsychENCODE^30^, which represent large human blood and brain gene expression datasets, respectively. We found no colocalization between PDD GWSS *loci* and eQTLs from either dataset, indicating that there is no evidence of shared causal variants driving both gene expression and the four association signals for PDD progression (Supplementary Table 4). Of note, *LRP1B* is not expressed in blood, thus no *LRP1B* eQTLs (significant or non-significant) were available from eQTLGen (Supplementary Fig. 5a). Next, we explored whether non-coding GWSS significant signals could have a role in alternative splicing by performing colocalization analysis using cortical and nigral splicing QTLs (sQTLs) from the Genotype-Tissue Expression (GTEx) dataset^31^. Again, we found no colocalization of PDD GWSS *loci* and sQTLs (Supplementary Table 5). We also generated regional associations plots for transcript expression QTLs (tQTL) from PsychENCODE and PDD GWSS signals in the region surrounding *LRP1B*, which upon visual assessment did not suggest the presence of signal colocalization between *LRP1B loci* and tQTLs (Supplementary Fig. 9). Despite the power limitations of existing QTL datasets, the available data does not currently support *LRP1B* signals regulating the expression of transcript isoforms via alternative splicing.

### APOE and LRP1B interaction

One of the ligands of *LRP1B* at the cell surface is *APOE*. To investigate whether the *LRP1B* signal was independent of *APOE* status, we defined four groups of PD patients in the combined cohorts (*n* = 3,694): non-carriers of either *APOE* ε4 or *LRP1B* rs80306347 alleles, exclusive carriers of *APOE* ε4 allele, exclusive carriers of *LRP1B* rs80306347 allele and carriers of both alleles. We then used a Cox proportional hazards (CPH) model in the combined cohorts to calculate the hazards of survival dementia-free in each of these groups, adjusting for sex, age at disease onset or diagnosis, the first five PCs and the cohort each individual originated from (Figure 2a). Compared to non-carriers, participants exclusively carrying the *LRP1B* rs80306347 allele had increased risk of progressing to PDD (HR = 2.19; 95% CI =1.23-3.88; *P* = 0.00730). In addition, we also performed survival analysis controlling for *APOE* status (Fig. 2b-c). An increased hazard of progression to PDD was present in *LRP1B* rs80306347 carriers in both *APOE* ε4 allele carriers (HR = 3.16; 95% CI = 1.78-5.61; *P* = 8.54x10^-05^) and *APOE* ε4 allele non-carriers (HR = 2.14; 95% CI = 1.21-3.82; *P* = 0.00947), confirming that the effect of rs80306347 is independent of the effect of *APOE*. Finally, individuals carrying both *APOE* ε4 and *LRP1B* rs80306347 alleles had a much higher hazard of progression to PDD than carriers of each allele separately (HR = 7.69; 95% CI = 4.42-13.4; *P* = 5.76x10^-13^), indicating increased risk of progression to PDD and possibly a functional interaction between the two alleles (Fig. 2a).

**Fig. 2.**
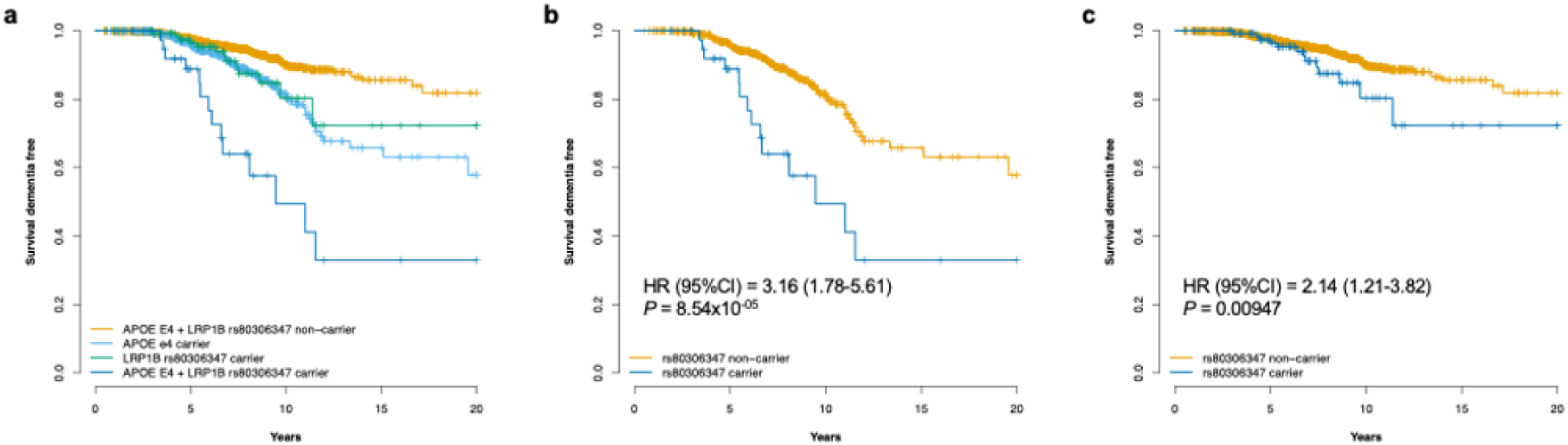
Interaction between *APOE* and *LRP1B* rs80306347 signals**. a** Kaplan-Meier curve for dementia-free survival based on *APOE* ɛ4 and *LRP1B* rs80306347 carrier status of PD patients. Compared to non-carriers of either allele, *LRP1B* rs80306347 carriers had an HR of progression to PDD of 2.19 (95% CI = 1.23-3.88; *P* = 0.00730), while *APOE* ɛ4 carriers had an HR of 2.43 (95% CI = 1.88-3.15; *P* = 1.40x10^-11^). Carriers of both alleles had the most significant increase in the hazards ratio of progressing to PDD (HR = 7.69; 95% CI = 4.42- 13.40; *P* = 5.76x10^-13^). **b** Kaplan-Meier curve for dementia-free survival based on *LRP1B* rs80306347 carrier status in PD *APOE* ɛ4 carriers. **c** Kaplan-Meier curve for dementia-free survival based on *LRP1B* rs80306347 carrier status in PD *APOE* ɛ4 non-carriers. Statistical analysis was conducted using Cox proportional hazards models in the combined cohorts (*n* = 3,964 individuals) at the specified *loci*. *HR*, hazard ratio; *CI*, confidence interval; *P*, *p-*value.

### Candidate gene analysis

Several other genes have been suggested to increase the risk of cognitive decline or dementia in PD. One of the most widely reported genes is *GBA*, which has also been described as a risk factor for PD and an earlier age of disease onset^32, 33^. The non-GD-causing *GBA* PD-risk variant E365K (rs2230288, previously known as E326K) has been described in association with cognitive progression in PD^12, 13^. We therefore performed a candidate *loci* survival analysis in the combined cohorts (*n* = 3,694) based on E365K carrier status, which confirmed a significant hazard ratio for progression to dementia (HR = 2.33; 95% CI = 1.50-3.61; *P* = 1.64 x10^-04^; Fig. 3a; Supplementary Table 6). In addition, *GBA* Sanger sequencing data was available for 1,793 individuals originating from the DIGPD and TPD cohorts. Mutations causing Gaucher’s disease and PD risk variants were combined for survival analysis and were present in 9.3% of the cases (Supplementary Table 7). In this subset of patients, *GBA* risk variant and GD-mutation carriers had a HR for progression to PDD of 2.02 (95% CI = 1.21-3.32; *P* = 0.007), confirming the observation from several previous studies that *GBA* mutations increase the risk of dementia (Supplementary Fig. 10)^11–13^. A similar candidate *loci* approach in the combined cohorts confirmed the strong association of *APOE* ε4 carrier status (HR = 2.54; 95% CI = 1.99-3.24; *P* = 8.16x10^-14^) and *LRP1B* rs80306347 carrier status (HR = 2.64; 95% CI = 1.76-3.95; *P* = 2.34x10^-06^) with earlier progression to PDD (Fig. 3b-c).

**Fig. 3.**
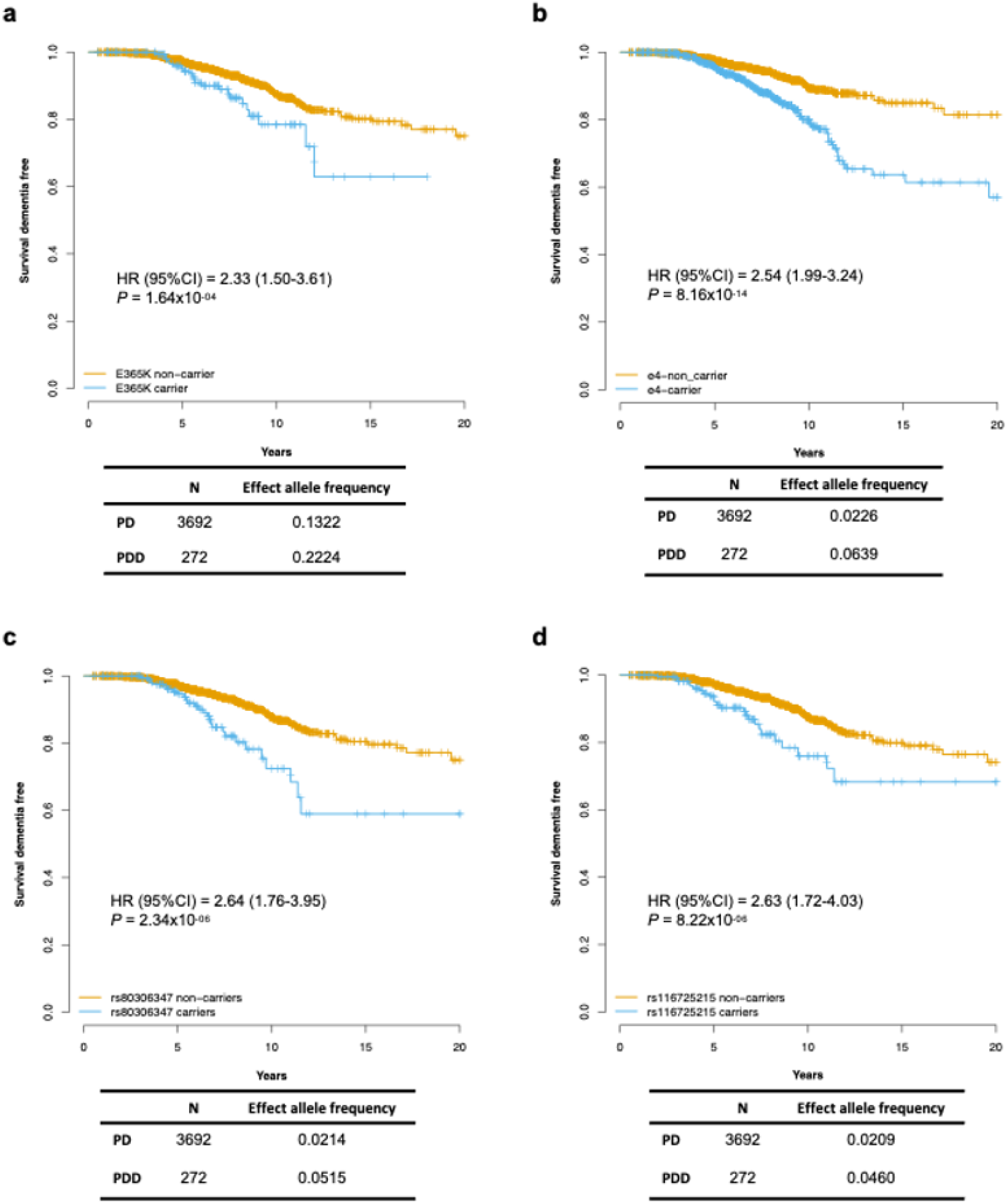
Survival curves of candidate gene analysis**. a** Kaplan-Meier curve for dementia-free survival based on *GBA* E365K carrier status of PD patients. **b** Kaplan-Meier curve for dementia-free survival based on *APOE* ɛ4 carrier status of PD patients. **c** Kaplan-Meier curve for dementia-free survival based on *LRP1B* rs80306347 carrier status of PD patients. **d** Kaplan-Meier curve for dementia-free survival based on *LRP1B* rs116725215 carrier status of PD patients. Statistical analysis was conducted per *locus* using Cox proportional hazards models in the combined cohorts (*n* = 3,964 individuals). *HR*, hazard ratio; *CI*, confidence interval; *P*, *p-*value.

Multiplications of *SNCA* can cause autosomal dominant PD that is often associated with a high prevalence of dementia^34^. In addition, common variants in *SNCA* have been reported to increase the risk of cognitive decline or dementia in PD patients, as well as the risk of DLB, a related parkinsonism disorder in which dementia is an early feature^35–37^. We investigated five *SNCA* variants previously reported in the literature for association with dementia in PD or DLB, but none were shown to increase the risk of progression to PDD (Supplementary Table 6). Some of these variants have only been reported in small studies^37^, while rs356219 has shown inconsistent results across studies^10, 38, 39^, indicating that there is not enough evidence to support a role for common *SNCA* variants in the risk of cognitive decline or dementia in PD. Importantly, variants identified in DLB case-control GWAS studies^36^ do not appear to contribute to risk of progression to dementia in PD, suggesting that the mechanisms leading to DLB and PDD do not entirely overlap.

Some studies have found that the *MAPT* H1 haplotype is a risk factor for cognitive decline in PD and can increase the susceptibility to DLB^9, 14, 15, 40^. However, this finding has not been consistently replicated^10, 41^. Similarly, we did not find any association between *MAPT* haplotypes and time to dementia in PD (Supplementary Table 6).

Recently, common variants in *RIMS2*, *TMEM108* and *WWOX* have been suggested to associate with faster progression to PDD^41^. Using similar methodology and sample size, we did not replicate these findings (Supplementary Table 6), indicating that further studies are needed to confirm the role of these genes in the risk of cognitive decline in PD.

### AD and PD genetic risk scores in PDD

Given the role of both APOE and LRP1B in APP metabolism, we next investigated the overlap between the AD risk profile with that of PD cases with and without dementia. We calculated the normalised individual-level genetic risk score in each of the cohorts, based on the summary statistics from a recent large-scale GWAS meta-analysis of Alzheimer’s disease^42^. A generalised linear model was used to test the association of AD genetic risk scores with dementia status in each cohort, with results further meta-analysed using a random-effects model (Fig. 4a and Supplementary Fig. 11a). PDD was associated with a higher genetic risk score for Alzheimer’s disease (Odds Ratio (OR) = 1.46, 95% CI = 1.31- 1.64, *P* = 7.22x10^-11^). In contrast, the normalised genetic risk score for Parkinson’s disease, derived from the latest PD GWAS study^32^, was similar between PDD and non-demented PD cases (OR = 1.0, 95% CI = 0.84-1.18, *P* = 0.9698) (Fig. 4b and Supplementary Fig. 11c). This suggests that the genetic risk of developing PDD overlaps with the risk of developing Alzheimer’s disease, potentially identifying APP metabolism as mechanistically relevant for the progression to dementia in the context of Parkinson’s disease. Interestingly, in a subset of PD samples from the AMP-PD cohort who have been tested for Alzheimer’s disease biomarkers in cerebrospinal fluid (CSF), PDD cases had decreased amyloid β (Aβ) 42 levels (median ± interquartile range (IQR): 581 ± 493 pg/mL vs 867 ± 478 pg/mL, *P* = 0.001193, Wilcoxon rank-sum test) and increased total Tau (208 ± 129 pg/mL vs 158 ± 70 pg/mL, *P* = 0.01617, Wilcoxon rank-sum test) and p-Tau181 (18.3 ± 14.3 pg/mL vs 13.3 ± 5.84 pg/mL, *P* = 0.002544, Wilcoxon rank-sum test) levels at baseline (Fig. 5a), supporting the hypothesis that APP metabolism is important for the development of PDD. In addition, *APOE* ε4 carriers also had significantly decreased CSF Aβ42 levels at baseline (median ± IQR: 689 ± 386 pg/mL vs 896 ± 543 pg/mL, *P* = 1.7x10^-06^, Wilcoxon rank-sum test) and subsequent time points, with no change in total Tau or p-Tau181 levels (Fig. 5b). This is in keeping with results from previous genome-wide association studies of AD biomarkers, showing an association of *APOE* with abnormal amyloid status in either CSF or PET scans^43–46^. *APOE* status is the most significant genetic determinant to the risk of developing Alzheimer’s disease^42^, and was also confirmed to be significantly associated with the risk of progression to PDD in individuals previously diagnosed with PD. Therefore, to establish that the association between Alzheimer’s disease genetic risk score and progression to PDD is not exclusively due to the overlap of the *APOE* signal between these two conditions, we adjusted the generalised linear models for *APOE* ε4 carrier status. When adjusting for *APOE* ε4 carrier status, there was no significant association between PDD and the genetic risk score for Alzheimer’s disease (OR = 1.20, 95% CI = 0.94-1.52, *P* = 0.1370), indicating that *APOE* ε4 carrier status alone is driving the risk of progression to dementia among AD GWAS top hits.

**Fig. 4.**
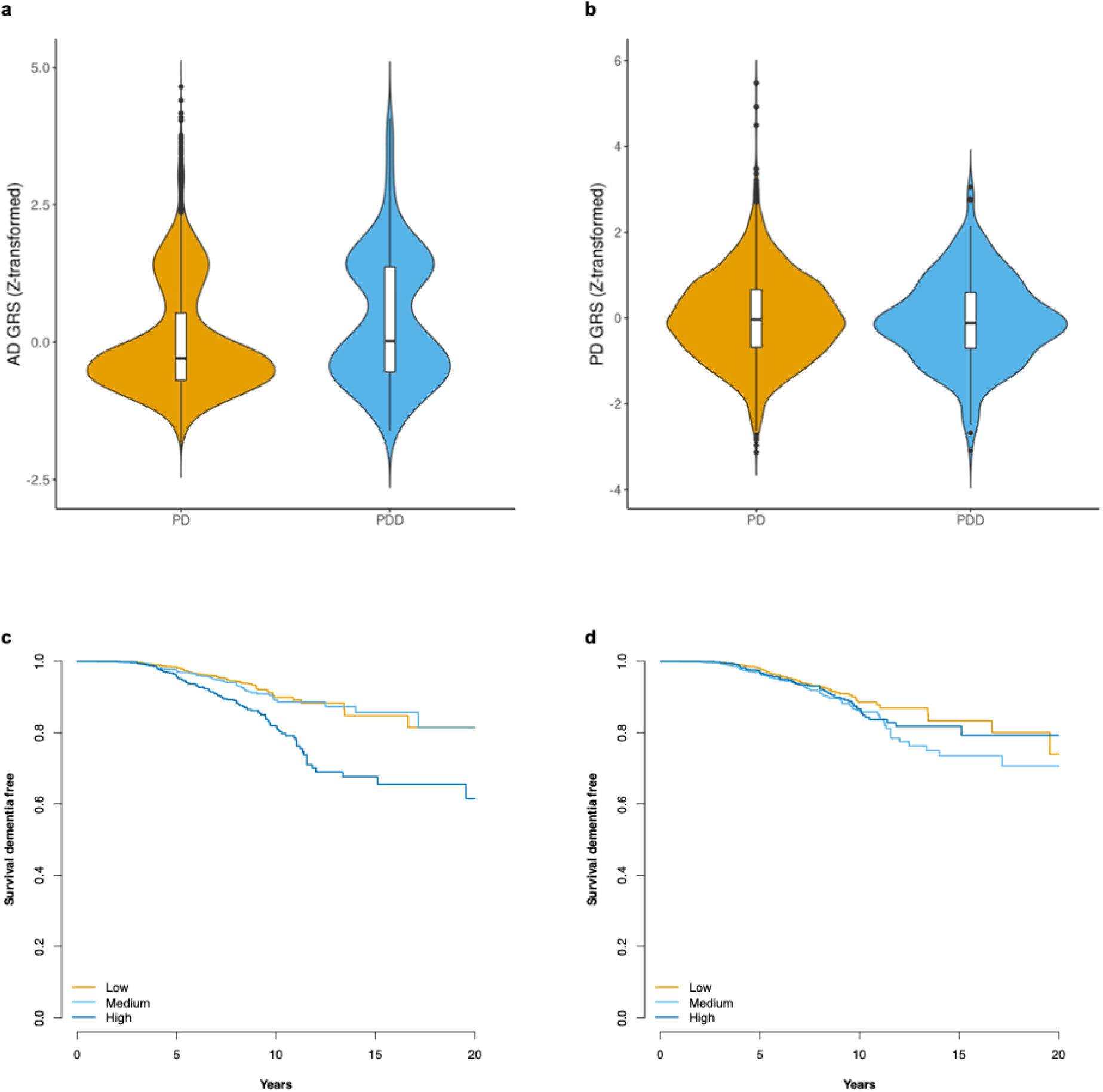
Alzheimer’s disease and Parkinson’s disease genetic risk scores (GRS). **a-b** Violin plots depicting the distribution of the meta-analysis of z-transformed Alzheimer’s disease (**a**) and PD genetic risk scores (**b**) in PD and PDD. The central line of the boxplots indicates the median, the box limits indicate the first and third quartiles, the whiskers indicate ±1.5*IQR, and the data points indicate the outliers. **c-d** Survival Kaplan-Meier curves for dementia-free survival of PD patients based on the stratification of AD-GRS into low-, middle-, and high-risk tertiles, either including (**c**) or excluding APOE (**d**).

**Fig. 5.**
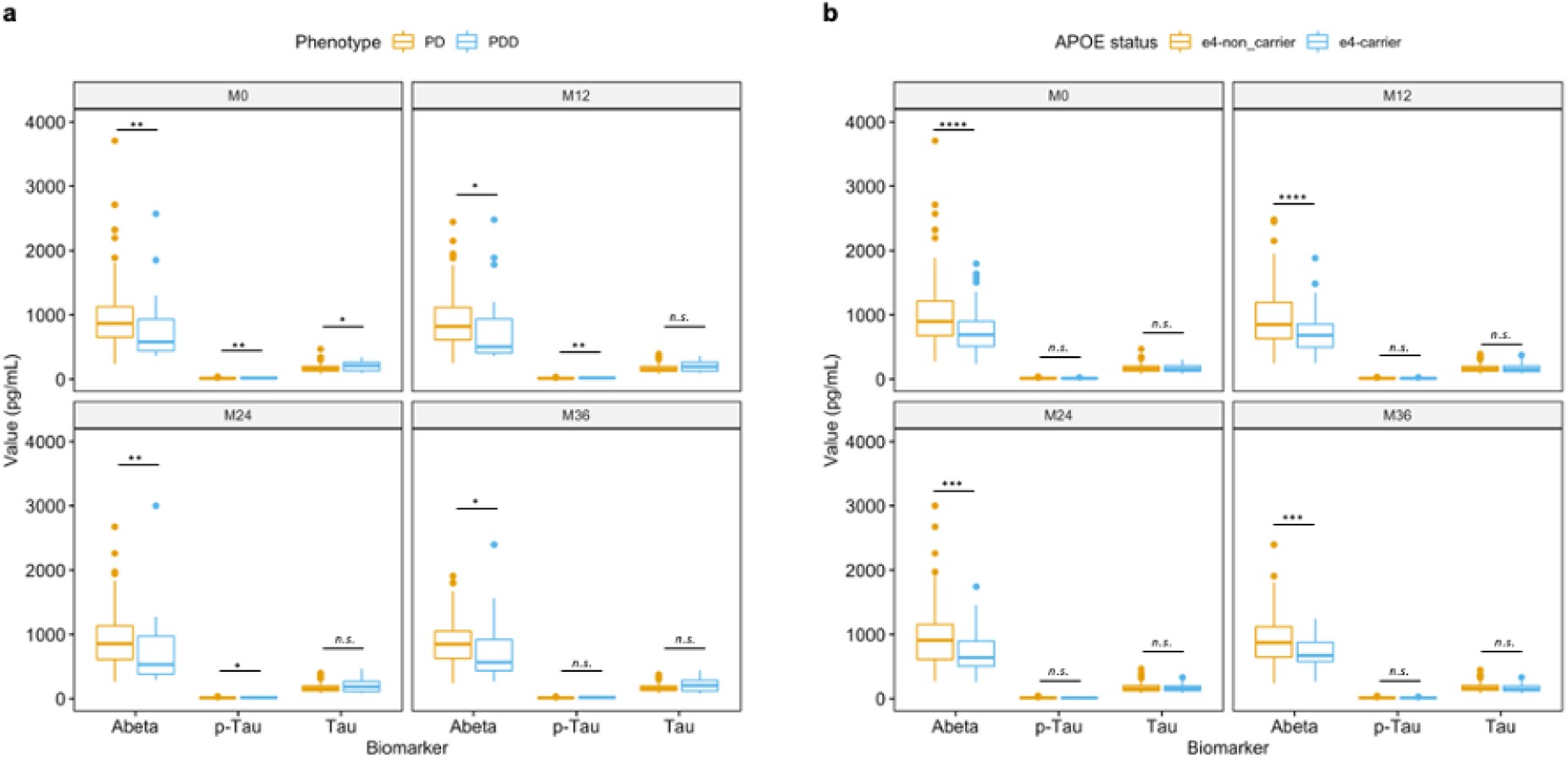
CSF measurements of AD biomarkers. Boxplots representing the measurements (in pg/mL) of the CSF biomarkers Aβ42, p-Tau181 and total Tau in a subset of individuals from the AMP-PD cohort (*n* = 352) across time (M0 = study baseline, M12 = 12 months, M24 = 24 months, M36 = 36 months). **a** CSF biomarker levels by phenotype (*n* = 28 PDD and *n* = 324 PD cases). **b** CSF biomarker levels by APOE Ɛ4 allele carrier status (*n* = 86 APOE Ɛ4 allele carriers and *n* = 266 APOE Ɛ4 allele non-carriers). Boxplots display a median line, the box limits indicate the first and third quartiles, the whiskers indicate ±1.5*IQR, and the data points indicate the outliers. The Wilcoxon rank-sum test was used to compare medians across phenotypic groups. Significance threshold: *P <0.05, **P <0.01, ***P <0.001, ****P <0.0001, n.s. not significant.

Finally, we asked whether a higher Alzheimer’s disease genetic risk score could be contributing to decreased dementia-free survival, i.e., faster progression to PDD. We performed survival analysis using CPH models to calculate the hazards of survival dementia-free after stratification of PD individuals into low-, middle-, and high-risk based on AD genetic risk scores. Individuals in the higher tertile of AD-GRS had faster progression to dementia (HR = 2.33, 95% CI = 1.72-3.14, *P* = 3.63x10^-08^), but as with the overall risk of PDD, faster progression to dementia was abolished after exclusion of the *APOE* signal (HR = 1.13, 95% CI = 0.83-1.54, *P* = 0.4418, Fig. 4c-d).

## Discussion

We have conducted a large genome-wide survival study of progression to dementia in PD patients. *APOE* has consistently been implicated as a risk factor for idiopathic PD, PDD and DLB^9, 19, 32, 35, 36, 47^. Our results confirm that *APOE* ε4 is also a significant contributing factor in the rate of progression to PDD, while a candidate gene approach confirmed the role of non-GD-pathogenic *GBA* E365K PD risk variant in accelerating cognitive decline in PD. In addition, we identified three novel *loci* associated with progression to dementia.

*LRP1B* belongs to the low-density lipoprotein (LDL) receptor family and is highly expressed in the brain^21^. Several members of the LDL family have been implicated in cellular processes relevant to neurodegeneration, including tau uptake^48^ and amyloid precursor protein (APP) trafficking, processing, and clearing^49^. Whether APP is processed by beta- and gamma-secretases to Aβ in the amyloidogenic pathway or by alpha-secretases in the non-amyloidogenic pathway depends on its subcellular localisation, as beta-secretase is most active in the acidic pH of the endosome, which appears to be a key site for the production of Aβ^50^. Therefore, modulation of intracellular APP tracking by LDL receptors with opposing activities is postulated to be a crucial determinant of APP processing and subsequent neurodegeneration^51^. For example, binding of LRP1 and LRAD3 to APP at the cell surface leads to its enhanced endocytic trafficking and increased processing to Aβ^52, 53^. In contrast, binding of LRP1B and LRP10 to APP leads to decreased trafficking of APP to the endosome, thus resulting in reduced amyloidogenic processing of APP^23, 54^. *LRP10* mutants that disrupt the distribution of LRP10 from the trans-Golgi network to early endosomes lead to increased presence of APP in the endosomes and consequently to increased amyloidogenic processing of APP^54^. Interestingly, loss of function mutations in *LRP10* have recently been implicated in familial PD^55^. Similarly, due to a slower rate of endocytosis which leads to APP accumulation at the cell surface, the binding of APP to LRP1B receptors reduces APP processing into Aβ and increases secretion of soluble APP instead, suggesting that enhanced LRP1B activity could protect against the pathogenesis of Alzheimer’s disease.^19^ Interestingly, a genome-wide study comparing elderly individuals without cognitive decline and those with late onset Alzheimer’s disease identified variants in *LRP1B* as protective against cognitive decline in old age^56^.

It is likely that dementia in PD can be driven by distinct mechanisms. Research in dementia with Lewy bodies, a condition closely related to PDD, has shown that *GBA* is more strongly associated with risk of “pure” DLB, while *APOE* ε4 is more strongly associated with DLB with AD co-pathology^57, 58^. This suggests that the genetic drivers of dementia in α- synucleinopathies are different in cases with and without Aβ co-pathology, with *GBA* predisposing to pure Lewy body pathology and *APOE* predisposing to concomitant Aβ deposition. While PD neuropathology is primarily characterised by deposition of α-synuclein aggregates, dementia in PD can also be associated with Aβ deposition^59–61^. This leads to the question of whether Aβ metabolism could also play an important role in the development of PDD. In fact, increased cortical Aβ deposition has been shown to be associated with a faster progression to dementia in PD^61, 62^, and a low CSF Aβ_42_-to-total tau ratio at baseline has been associated with cognitive decline in early PD^63^. Our results on AD CSF biomarkers also suggest that PDD is associated with increased Aβ brain pathology, and it is likely that *APOE* ε4 is the main driver of this association. Furthermore, APOE is known to facilitate endocytosis of Aβ via LDL receptors at the cell surface^64^, which could offer a mechanistic link between *APOE* and *LRP1B* and a possible explanation as to why PD carriers of both *APOE* ε4 and *LRP1B* rs80306347-C alleles appear to have faster progression to dementia. Nonetheless, there is evidence that *APOE* ε4 can also contribute to neurodegeneration by non-amyloidogenic mechanisms: *APOE* ε4 allele carriers can present with “pure” Lewy body dementia (LBD); α-synuclein pathology is increased in LBD *APOE* ε4 carriers with minimal amyloid pathology, compared to age-matched non-carriers; *APOE* ε4 exacerbates α- synuclein pathology and leads to worse neurodegeneration and cognitive performances in mice^65, 66^.

We have identified *SLC6A3* and *SSR1* as promising novel candidates associated with progression to PD dementia. *SLC6A3* encodes the dopamine transporter DAT and is essential in the regulation of dopamine metabolism and neurotransmission. Dopaminergic deficits have been suggested as contributing to cognitive impairment in PD^67^, highlighting the potential of *SLC6A3* to influence cognitive performance in these patients. Polymorphisms in *SLC6A3* have been shown to have a functional impact on dopamine metabolism and responses to exogenous dopamine^68^, which could indicate that the effect of *SLC6A3* on cognition could be mediated by changes in dopamine reuptake that would affect both endogenous and therapeutic dopamine. Since only one of the studies in the meta-analysis was found to be driving the signal at this locus, more research is needed to support a role for *SLC6A3* in the progression to PDD. Finally, the variant rs60707664 on chromosome 6p24.3 is located 7.5 kb upstream of the *SSR1* gene. *SSR1* encodes the translocon-associated protein subunit alpha, a subunit of the signal sequence receptor (SSR), a glycosylated ER membrane receptor associated with protein translocation across the ER membrane. ER dysfunction leads to the accumulation of misfolded proteins and triggers ER stress, which in turn activates the unfolded protein response (UPR) that has been shown to play a role in PD pathogenesis^27^. Remarkably, *SSR1* expression has recently been shown to be upregulated in an early PD mouse model and to be highly correlated with the loss of TH neurons, making *SSR1* a plausible candidate for neurodegeneration in PD^69^.

In conclusion, this large genome-wide study identifies several interesting and plausible new gene candidates associated with faster progression to dementia in PD, while also corroborating the importance of the previously described *APOE* and *GBA* variants for cognitive outcomes in PD. In addition, we provide further evidence for the role of β-amyloid metabolism in the development of dementia in PD, which has important therapeutic implications, as strategies aimed at Alzheimer’s disease could also prove effective in PD patients at risk of dementia.

## Methods

### Patient cohorts

We have studied four independent longitudinal PD cohorts: Tracking Parkinson’s (TPD)^16^, Oxford Parkinson’s Disease Centre Discovery Cohort (OPDC)^17^, Accelerating Medicines Partnership: Parkinson’s Disease (AMP-PD, v2.5), which consists of harmonised data from multiple cohorts^18^, and Drug Interaction With Genes in Parkinson’s Disease (DIGPD), comprising a total of 3,964 participants after clinical and genetic data cleaning (Supplementary Fig. 1 and Supplementary Table 1). All participants were diagnosed with PD according to the Queen Square Brain Bank criteria^70^. Participants were excluded from the analysis if an alternative diagnosis was made during the follow-up period (including a diagnosis of DLB) and/or the probability of a PD diagnosis as assessed by a clinician at the last available visit was less than 90%. In AMP-PD, only individuals in the PD study arm were included. Criteria for PDD was based on the Movement Disorders Society (MDS) taskforce PDD diagnostic criteria^3, 41^. Specifically, participants were classified as having PDD if they had adjusted MoCA scores < 21/30, at least two cognitive domains impaired in the MoCA scale (attention/serial sevens ≤ 2/3; language / verbal fluency 0/1; memory / delayed recall ≤ 4/5; visuospatial/executive ≤ 4/5), a cognitive deficit severe enough to impact on activities of daily living (MDS-Unified PD Rating Scale (UPDRS) part I 1.1 ≥ 2 score), and absence of severe depression (MDS-UPDRS part I 1.3 < 4), except participants from the DIGPD cohort, for whom only MMSE scores were available together with a clinician assigned diagnosis of dementia. Participants were excluded from the study (left censored) if they met criteria for PDD at study baseline (Supplementary Table 8).

Time-to-event was calculated as the number of years from disease onset (or disease diagnosis, in the case of the AMP-PD cohort) until the first visit where criteria for PDD was met or until study withdrawal due to dementia. Individuals with missing data regarding time-to-event or event classification were also excluded from the study. Comparisons across cohorts were performed in R (R Project for Statistical Computing, RRID:SCR_001905; version 4.1.3; https://www.R-project.org/) using Chi-squared test with Yates’ continuity correction (*rstatix* package, version 0.7.0; RRID:SCR_021240; https://CRAN.R-project.org/package=rstatix) for categorical variables, and Kruskal-Wallis test with Dunn’s test for post hoc multiple pairwise comparisons (*stats* package, version 4.1.3; https://stat.ethz.ch/R-manual/R-devel/library/stats/html/00Index.html) for continuous variables, with p-values adjusted by the Bonferroni method. Significance was set at α=0.05.

### Data Quality Control

Whole-genome sequence data was available from participants in AMP-PD cohorts. The remainder of samples were genotyped with the Illumina HumanCoreExome array (TPD), Illumina HumanCoreExome-12 v1.1 or Illumina Infinium HumanCoreExome-24 v1.1 arrays (OPDC) and the Illumina Infinium Multi-Ethnic Global (MEGA) array (DIGPD). Sample quality control (QC) included the exclusion of samples with call rates <98%, samples with excess heterozygosity (defined as samples deviating more than two standard deviations from the mean heterozygosity rate), samples with a mismatch between clinical sex and genetically determined sex from chromosome X heterogeneity, and samples from related individuals (pi- hat > 0.125). Variants with missingness rate >5%, minor allele frequency (MAF) <0.01 and Hardy-Weinberg equilibrium (HWE) *P* <1x10^-08^ were excluded. We opted for a more liberal HWE threshold given the case-case study design and to account for the fact that disease-associated variants which may deviate from HWE could also be associated with disease progression. To identify the ancestry, variants in linkage disequilibrium were removed and samples clustered against the HapMap3 reference panel, using principal component analysis. Individuals who deviated more than 6 standard deviations from the mean of the first 10 principal components of the HapMap3 CEU+TSI population were excluded from the analysis (Supplementary Fig. 12a). After extraction of European-ancestry samples, principal components were re-calculated in the study data to use as covariates. The genotyping array data was then imputed against the Haplotype Reference Consortium (HRC) reference panel (version r1.1 2016; http://www.haplotype-reference-consortium.org/) in the Michigan Imputation Server (RRID:SCR_017579; https://imputationserver.sph.umich.edu)^71^ using Minimac4 (version 1.0.0; https://genome.sph.umich.edu/wiki/Minimac4 version 1.0.0). Imputed variants were excluded if the imputation info R^2^ score was ≤ 0.3. Following imputation, variants with missingness > 5%, minor allele frequencies <1% and in extreme Hardy-Weinberg disequilibrium (*P* < 1x10^-08^) were also excluded. Data cleaning was performed using PLINK v1.9 (RRID:SCR_001757; https://www.cog-genomics.org/plink/1.9/)^72^.

### Time-to-event GWSS and meta-analysis

A time-to-event genome-wide survival study (GWSS) was performed in R (version 4.1.2) in each cohort, using the Cox proportional hazards function in the *survival* package (version 3.2.13; RRID:SCR_021137; https://CRAN.R-project.org/package=survival), in which time to PDD was regressed against each SNP, with age at onset or diagnosis, sex and first five principal components as covariates. AMP-PD summary statistics were converted from hg38 to hg19 using the binary liftOver tool (RRID:SCR_018160; https://genome.sph.umich.edu/wiki/LiftOver). The summary results from each cohort were then meta-analysed using METAL software in a random effects model, using genomic control correction (version released on the 25/03/2011; RRID:SCR_002013; http://csg.sph.umich.edu//abecasis/Metal/)^73^. The genomic inflation factor (λgc) for each cohort varied between 0.710 and 1.003. After the meta-analysis, the λgc was 1.048 (Supplementary Fig. 12b). Upon completion of the meta-analysis, variants that were not present in all samples were excluded, as well as variants with minor allele frequency variability >15% across studies. Variants were also excluded if the p-value for the Cochran’s Q-test for heterogeneity was less than 0.05 and the I^2^ statistic was ≤80%.

Forest plots of variants of interest were prepared using the R package *forestplot* (version 2.0.1; https://CRAN.R-project.org/package=forestplot). Results of the meta-analysis were annotated using FUMA (Functional Mapping and Annotation of Genome Wide Association Studies, RRID:SCR_017521; version 1.3.8; https://fuma.ctglab.nl/)^16^. Regional association plots were generated in LocusZoom (RRID:SCR_021374; http://locuszoom.org/)^74^. LDproxy (version 5.2; https://ldlink.nci.nih.gov/?tab=ldproxy)^75^ was used to identify variants in high linkage disequilibrium with variants of interest.

### Tissue and cell-type specificity measures

Specificity represents the proportion of a gene’s total expression attributable to one cell type/tissue. To determine specificity of a gene to a tissue or cell-type, specificity values from three independent gene expression datasets were generated. Briefly, these datasets included 1) bulk-tissue RNA-sequencing of 53 human tissues from the Genotype-Tissue Expression consortium (GTEx, version 8; RRID:SCR_013042)^31^; 2) human single-nucleus RNA-sequencing of the middle temporal gyrus from the Allen Institute for Brain Science (AIBS, Allen Cell Types Database - Human MTG Smart-Seq 2018 dataset, available from celltypes.brain-map.org/rnaseq; RRID:SCR_014806)^76^; and 3) human single-nucleus RNA-sequencing of the substantia nigra^77^. Generation of specificity values for GTEx and AIBS were previously described in Chia *et al* ^35^. Briefly, specificity values for GTEx were generated using code modified from a previous publication (https://github.com/jbryois/scRNA_disease)^78^, to reduce redundancy among brain regions and to include protein- and non-protein-coding genes. Specificity values for the AIBS-derived dataset were generated using gene-level exonic reads and the ‘generate.celltype.data’ function of the *EWCE* R package (version 1.2.0)^79^. Likewise, specificity values from Agarwal *et al.* were generated using *EWCE*. Specificity values for all three datasets and the code used to generate these values are openly available at https://github.com/RHReynolds/MarkerGenes^80^.

### Colocalization analysis

To investigate whether there is an overlap between PDD *loci* and expression quantitative trait *loci* (eQTLs), we used the *coloc* R package (version 5.1.0; https://cran.r-project.org/web/packages/colocr/index.html)^81^. We also used the R package *colochelpR* (version 0.99.0)^82^ to help prepare datasets for use with *coloc*. We took a Bayesian inference approach to test the H4 null hypothesis that there is a shared causal variant associated with both progression to PDD and gene expression regulation. The Bayesian inference approach additionally computes the posterior probability (PP) that there is no association with either trait (H0), there is association with the PDD trait but not the eQTL trait (H1), there is association with the eQTL trait but not the PDD trait (H2), and that there is association with both traits, but the causal variants are independent (H3). We extracted all the genes within 1 Mb of each significant *locus* in the PDD GWSS. *Coloc* was run using default p_1_=10^−4^, p_2_=10^−4^, and p_12_=10^−5^ priors (p_1_ and p_2_ are the prior probability that any random SNP in the region is associated with trait 1 and 2, respectively, while p_12_ is the prior probability that any random SNP in the region is associated with both traits). A PPH4 > 0.9 was considered evidence for the presence of a shared variant between traits, i.e., signal colocalization. Cis-eQTL data were obtained from 1) eQTLGen, comprising bulk blood-derived gene expression from 31,684 individuals (https://www.eqtlgen.org/cis-eqtls.html, accessed on the 07/06/2021) and 2) PsychENCODE, comprising gene expression from bulk RNA sequencing from the prefrontal cortex of 1,387 individuals (http://resource.psychencode.org/, accessed on the 07/06/2021)^29, 83^. Next, to understand if *LRP1B*, *SSR1* or *SLC6A3 loci* regulate alternative splicing, we used a similar approach using frontal cortex and substantia nigra sQTL data from the GTEx v8 database containing all variant-gene associations from 183 and 100 individuals, respectively, based on LeafCutter (version 0.2.9; RRID:SCR_017639; https://davidaknowles.github.io/leafcutter/)^84^ intron excision phenotypes. For *LRP1B*, we tested the alternative splicing from 8 different introns. In addition, FDR-filtered transcript-per-million (TPM) tQTLs (FDR <0.001) were obtained from PsychENCODE and used to generate regional association plots overlapping with *LRP1B* signals. A full colocalization analysis for tQTLs was not possible due to the unavailability of unfiltered tQTL summary statistics from PsychENCODE.

### Conditional analysis

To understand if one or more genome-wide significant variants at the same *locus* were contributing to the signal, we performed conditional analysis on single SNPs using a conditional and joint association analysis approach. We used the GWSS meta-analysis summary statistics and the entire AMP-PD cohort (*n* = 10,418) as the reference sample for linkage disequilibrium estimation. The reference sample was subjected to the same QC steps as described above. We then used CGTA-COJO software (version 1.93.0 beta for Linux; https://yanglab.westlake.edu.cn/software/gcta/#Overview)^85^ to perform association analysis conditional on SNPs of interest.

### Signal interaction between APOE and LRP1B

Given the affinity of LRP1B for ApoE carrying lipoproteins, we conducted a survival analysis based on *APOE* ε4 haplotype and *LRP1B* rs80306347 carrier status to understand if the effect of *LRP1B* rs80306347 signal was dependant on *APOE*. *APOE* genotypes were inferred from the imputed genotypes of rs7412 and rs429358 variants. Participants of the combined cohorts (*n* = 3,964) were grouped according to the presence of the two markers either simultaneously or exclusively, and a Cox proportional hazards model adjusted for age at onset or diagnosis, gender, the first five principal components and a cohort term was performed. We also conditioned the analysis on *APOE* ε4 carrier status by performing a survival analysis of *LRP1B* rs80306347 on *APOE* ε4 carriers and non-carriers separately.

### Candidate loci analysis

We additionally performed a candidate *loci* analysis of specific *loci* or variants of interest in the combined cohorts to increase power (*n* = 3,964), using Cox proportional hazards models adjusted for age at onset or diagnosis, sex, the first five principal components and a cohort term. The regions of interest consisted of genetic variants or *loci* previously identified in association with cognitive impairment in PD and/or DLB: *APOE* ε4 haplotype (rs429358)^8–10^, *GBA* E365K variant (rs2230288)^11, 12^, *SNCA* (rs356219, rs7680557, rs7681440, rs11931074, rs7684318)^35–37, 86^, *MAPT* H1 haplotype (rs1800547)^9, 14, 15^, *RIMS2* (rs182987047), *TMEM108* (rs138073281), and *WWOX* (rs8050111)^41^. In addition, participants from DIGPD and a subset of individuals from the TPD study were Sanger sequenced for GBA (*n* = 1,793). We performed a survival analysis as above based on GBA carrier status, for which we defined *GBA* mutation carriers as individuals with at least one Gaucher disease (GD)-causing mutation or PD risk factor (Supplementary Table 7).

### Genetic risk scores

To understand if there is overlap in the risk of development of PDD and the risk of PD or AD, we performed a genetic risk score (GRS) analysis using PLINK v1.9 software^72^. Scores were calculated using the summary statistics from the largest PD GWAS to date and the 2019 genome-wide association meta-analysis of AD, respectively^32, 42^. Only the independent genome-wide significant risk signals were used in the analysis. Scores were then z-transformed and added as a covariate in a logistic regression model, together with age at onset, sex and the first five PCs. Each cohort was analysed independently, and results were meta-analysed using the *meta* R package (version 5.1-1; RRID:SCR_019055; https://CRAN.R-project.org/package=meta)^87^. We conducted the AD-GRS analysis also without the *APOE* signal to investigate if the effect of AD-GRS in the risk of developing PDD was mediated by factors independent of *APOE*. For the survival analysis based on AD-GRS, individuals were stratified into low-, middle-, and high-risk tertiles of raw AD-GRS. We used Cox proportional hazards models adjusted for age at onset, sex and the first five PCs in each cohort; results were then meta-analysed with the R package *meta*.

### Association of clinical phenotype and APOE genotype with CSF biomarkers

A subset of AMP-PD participants (from the BioFIND and PPMI studies) included in the analysis have longitudinal cerebrospinal fluid (CSF) AD biomarker data available (*n* = 434). We investigated the association of phenotype (PDD vs PD) and *APOE* ε4 carrier status with average levels of amyloid β 42 (Aβ42), total Tau and tau phosphorylated at threonine 181 (p- Tau181) using unpaired two-sample Wilcoxon rank-sum tests (R *stats* package, version 4.1.2) at baseline, 12, 24 and 36 months of follow-up. Significance was set at α=0.05.

### Statistical power modelling

The R package *survSNP* (https://cran.r-project.org/web/packages/survSNP/index.html; version 0.25)^88^ was used to model statistical power for a hypothetical SNP with minor allele frequency of 2% and a hazards ratio of 2. The time-to-event was fixed at 4.5 years. Modelling took into account the event rates observed in the different cohorts.

## Data availability

Meta-analysis summary statistics are available for download from https://pdgenetics.org/resources. TPD data is available upon access request from https://www.trackingparkinsons.org.uk/about-1/data/. BioFIND, PPMI, PDBP and SURE-PD3 cohorts were accessed from Accelerating Medicines Partnership: Parkinson’s Disease (AMP-PD) and data is available upon registration at https://www.amp-pd.org/. OPDC data is available upon request from the Dementias Platform UK (https://portal.dementiasplatform.uk/Apply). DIGPD data is available upon request to the principal investigator (JC Corvol, Assistance Publique Hôpitaux de Paris). HapMap phase 3 data (HapMap3) is available for download at ftp://ftp.ncbi.nlm.nih.gov/hapmap/. The Ashkenazi Jewish population panel is accessible at https://www.ncbi.nlm.nih.gov/gds (accession ID: GSE23636). Cis-QTL data were obtained from eQTLGen (https://www.eqtlgen.org/cis-eqtls.html) and PsychENCODE (http://resource.psychencode.org). FDR-filtered tQTL data was obtained from PsychENCODE (http://resource.psychencode.org/). Cortical sQTL data was accessed from the GTEx v8 database (https://gtexportal.org/home/). GTEx bulk-tissue RNA-seq data is available at https://www.gtexportal.org/home/datasets. AIBS human single-nucleus RNA-seq data is available at https://portal.brain-map.org/atlases-and-data/rnaseq. Human single-nucleus RNA-seq of the substantia nigra data can be accessed from https://www.ncbi.nlm.nih.gov/geo/ (accession ID: GSE140231). Summary statistics from PD GWAS (Nalls et al, 2019) used to perform GRS analysis available from https://pdgenetics.org/resources.

## Code availability

Code used in the analysis is available from https://github.com/huw-morris-lab/PDD_GWSS (doi:<u>10.5281/zenodo.6535455</u>). Analysis was performed using open-source tools as described in the Methods section.

## Data Availability

All data produced in the present study are available upon request to the authors. Code used in the analysis is available online at https://github.com/huw-morris-lab/PDD_GWSS.

## Acknowledgements

This research was funded in whole or in part by Aligning Science Across Parkinson’s [Grant number: ASAP-000478] through the Michael J. Fox Foundation for Parkinson’s Research (MJFF). For the purpose of open access, the author has applied a CC BY public copyright licence to all Author Accepted Manuscripts arising from this submission.

This research was supported by the National Institute for Health Research University College London Hospitals Biomedical Research Centre. The UCL Movement Disorders Centre is supported by the Edmond J. Safra Philanthropic Foundation.

Data used in the preparation of this article were obtained from the AMP-PD Knowledge Platform (https://www.amp-pd.org). AMP-PD is a public-private partnership managed by the FNIH and funded by Celgene, GSK, Michael J. Fox Foundation for Parkinson’s Research, the National Institute of Neurological Disorders and Stroke, Pfizer, and Verily.

Clinical data and biosamples used in preparation of this article were obtained from the Fox Investigation for New Discovery of Biomarkers (BioFIND), the Parkinson’s Progression Markers Initiative (PPMI), the Parkinson’s Disease Biomarkers Program (PDBP), and the SURE-PD3 Study. BioFIND is sponsored by The Michael J. Fox Foundation for Parkinson’s Research (MJFF) with support from the National Institute for Neurological Disorders and Stroke (NINDS).

Data used in the preparation of this article were obtained from the Fox Investigation for New Discovery of Biomarkers (“BioFIND”) database (http://biofind.loni.usc.edu/). For up-to-date information on the study, visit michaeljfox.org/news/biofind.

PPMI – a public-private partnership – is funded by the Michael J. Fox Foundation for Parkinson’s Research and funding partners, including [list the full names of all of the PPMI funding partners found at www.ppmi-info.org/fundingpartners]. The PPMI Investigators have not participated in reviewing the data analysis or content of the manuscript. For up-to-date information on the study, visit www.ppmi-info.org.

The Parkinson’s Disease Biomarker Program (PDBP) consortium is supported by the National Institute of Neurological Disorders and Stroke (NINDS) at the National Institutes of Health. A full list of PDBP investigators can be found at https://pdbp.ninds.nih.gov/policy. The PDBP Investigators have not participated in reviewing the data analysis or content of the manuscript.

The DIGPD cohort (ClinicalTrials.gov, NCT01564992) is a multicenter longitudinal cohort conducted in four Universities and four General Hospitals in France between 2009 and 2019, sponsored by Assistance Publique Hôpitaux de Paris, funded by a grant from the French Ministry of Health (PHRC 2008, AOR0810) and a grant from the Agence Nationale de Sécurité et des Médicaments (ANSM-2013). We thank the DIGPD Study group which collected the data made available for this work.

Both TPD and OPDC cohorts are primarily funded and supported by Parkinson’s UK (https://www.parkinsons.org.uk/) and supported by the National Institute for Health Research (NIHR) Dementias and Neurodegenerative Diseases Research Network (DeNDRoN). The TPD study is also supported by NHS Greater Glasgow and Clyde. The OPDC cohort is also supported by the NIHR Oxford Biomedical Research Centre, based at the Oxford University Hospitals NHS Trust, and the University of Oxford. TPD has multi-centre research ethics approval from the West of Scotland Research Ethics Committee: IRAS 70980, MREC 11/AL/0163 (ClinicalTrials.gov, NCT02881099). OPDC has multi-centre research ethics approval from the South Central Oxford A Research Ethics Committee 16/SC/0108. Each subject provided written informed consent for participation.

The Genotype-Tissue Expression (GTEx) Project was supported by the Common Fund of the Office of the Director of the National Institutes of Health, and by NCI, NHGRI, NHLBI, NIDA, NIMH, and NINDS. The data used for the analyses described in this manuscript were obtained from https://console.cloud.google.com/storage/browser/gtex-resources on 01/26/2022.

## Author Contributions

H.R.M. and R.R. designed the study. H.R.M. supervised the study. D.G.G., M.T.M.H., Y.B.- S., J.H. and H.R.M. conceived and led the TPD and OPDC clinical cohorts, as well as performed data management and curation. J.C.C., A.B., S.L and J.F. conceived and led the DIGPD clinical cohort, as well as performed data management and curation. C.B. and L.H. performed genotyping and GBA Sanger sequencing of TPD samples. R.R. interrogated all the clinical data for the current study and performed quality control of AMP-PD genomic data. M.M.X.T. performed quality control of TPD, OPDC and DIGPD genomic data. R.R. performed the genome-wide survival analysis, candidate gene analysis, genetic risk score analysis and biomarker analysis. A.C.M. performed the colocalization analysis, with the contribution of R.H.R. R.H.R. wrote the pipeline for bulk tissue and single cell specificity. M.S. wrote the pipeline for the genome-wide survival analysis. M.R. consulted on the colocalization analysis. R.R. wrote the initial manuscript. All authors critically reviewed the manuscript.

## Competing Interests statement

H.R.M. reports paid consultancy from Roche and research grants from Parkinson’s UK, Cure Parkinson’s Trust, PSP Association, CBD Solutions, Drake Foundation, Medical Research Council (MRC), Michael J. Fox Foundation. Dr Morris is a co-applicant on a patent application related to C9ORF72 - Method for diagnosing a neurodegenerative disease (PCT/GB2012/052140).

D.G.G. has received grants from Michael’s Movers, the Neurosciences Foundation, and Parkinson’s UK, and honoraria from AbbVie, BIAL Pharma, Britannia Pharmaceuticals, GE Healthcare, and consultancy fees from Acorda Therapeutics and the Glasgow Memory Clinic.

M.T.M.H. received funding/grant support from Parkinson’s UK, Oxford NIHR BRC, University of Oxford, CPT, Lab10X, NIHR, Michael J. Fox Foundation, H2020 European Union, GE Healthcare and the PSP Association. She also received payment for Advisory Board attendance/consultancy for Biogen, Roche, Sanofi, CuraSen Therapeutics, Evidera, Manus Neurodynamica, Lundbeck.

Y.B.-S. has received grant funding from the MRC, NIHR, Parkinson’s UK, NIH, and ESRC. J.C.C. has served on advisory boards for Biogen, Denali, Idorsia, Prevail Therapeutic, Servier, Theranexus, UCB, and received grants from Sanofi and the Michael J. Fox Foundation outside of this work.

A.E. received funding/grant support by Agence Nationale de la Recherche, France Parkinson, and the Michael J. Fox foundation.

J.H. is supported by the UK Dementia Research Institute, which receives its funding from DRI Ltd, funded by the UK Medical Research Council, Alzheimer’s Society, and Alzheimer’s Research UK. He is also supported by the MRC, Wellcome Trust, Dolby Family Fund, National Institute for Health Research University College London Hospitals Biomedical Research Centre.

All other authors report no competing interests.

## Notes

### Competing Interest Statement

The authors have declared no competing interest.

### Clinical Protocols

https://github.com/huw-morris-lab/PDD_GWSS

### Author Declarations

TPD has multi-centre research ethics approval from the West of Scotland Research Ethics Committee (REC reference: 11/AL/0163). OPDC has multi-centre research ethics approval from the South Central Oxford A Research Ethics Committee (REC reference: 16/SC/0108). AMP-PD clinical data collection and DNA samples were obtained with local institutional and ethical approvals (details can be obtained from the https://amp-pd.org and each study website). DIGPD was sponsored by Assistance Publique Hopitaux de Paris and approved by French regulatory authorities (NCT01564992). All subjects provided written informed consent.

