## Supplementary Information for "Association between the *LRP1B* and *APOE loci* and the development of Parkinson’s disease dementia"

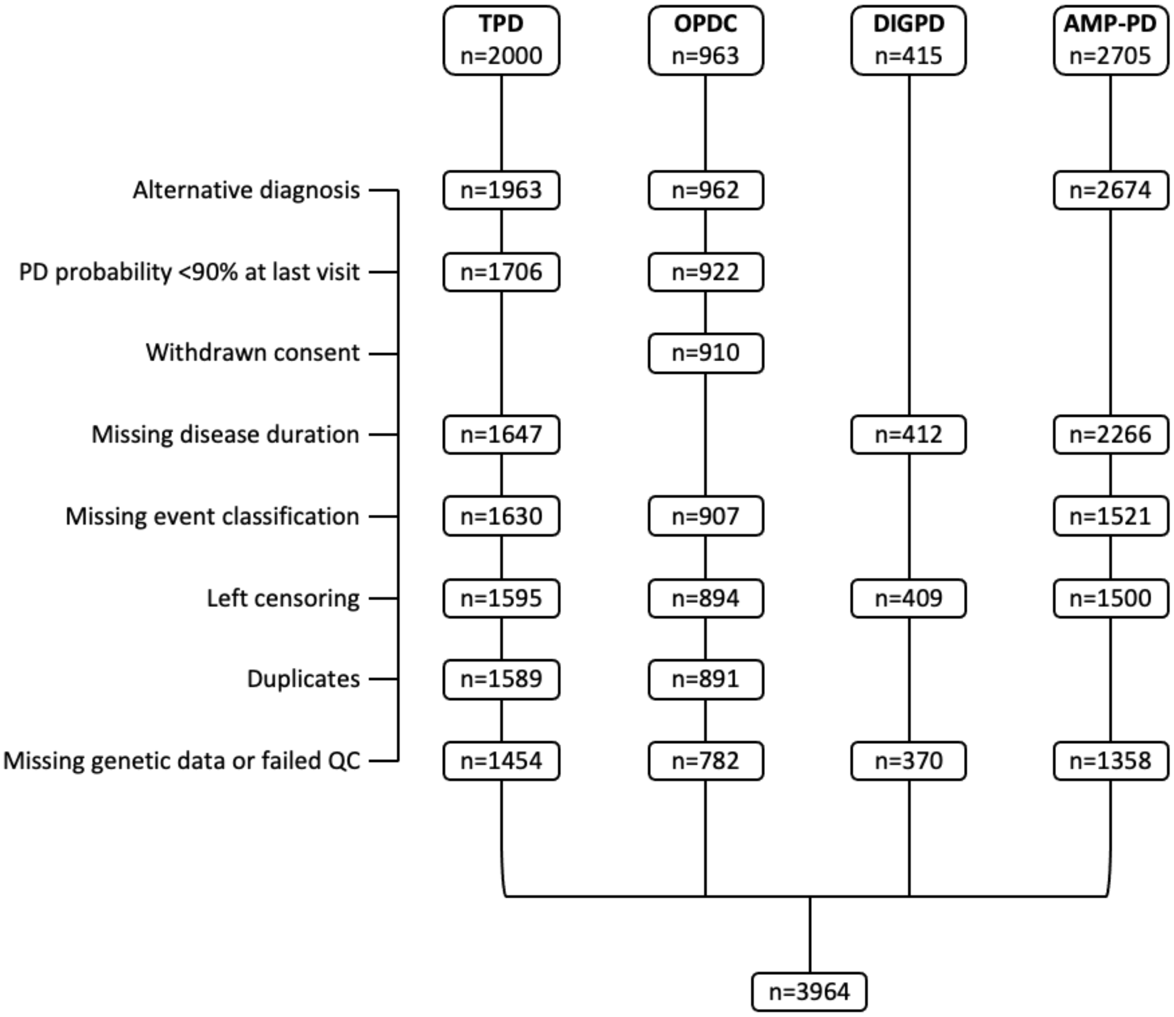


**Supplementary Fig. 1: Schematic of data cleaning workflow.**
Disease duration is time from disease onset or diagnosis until endpoint (dementia or censoring). Event classification is based on MoCA and UPDRS-I scores according to modified MDS Task Force criteria for PDD, except in the DIGPD cohort where it is based on clinician-assigned diagnosis of dementia. Left censoring refers to participants who had dementia at baseline and so could not be included in time to event analysis.

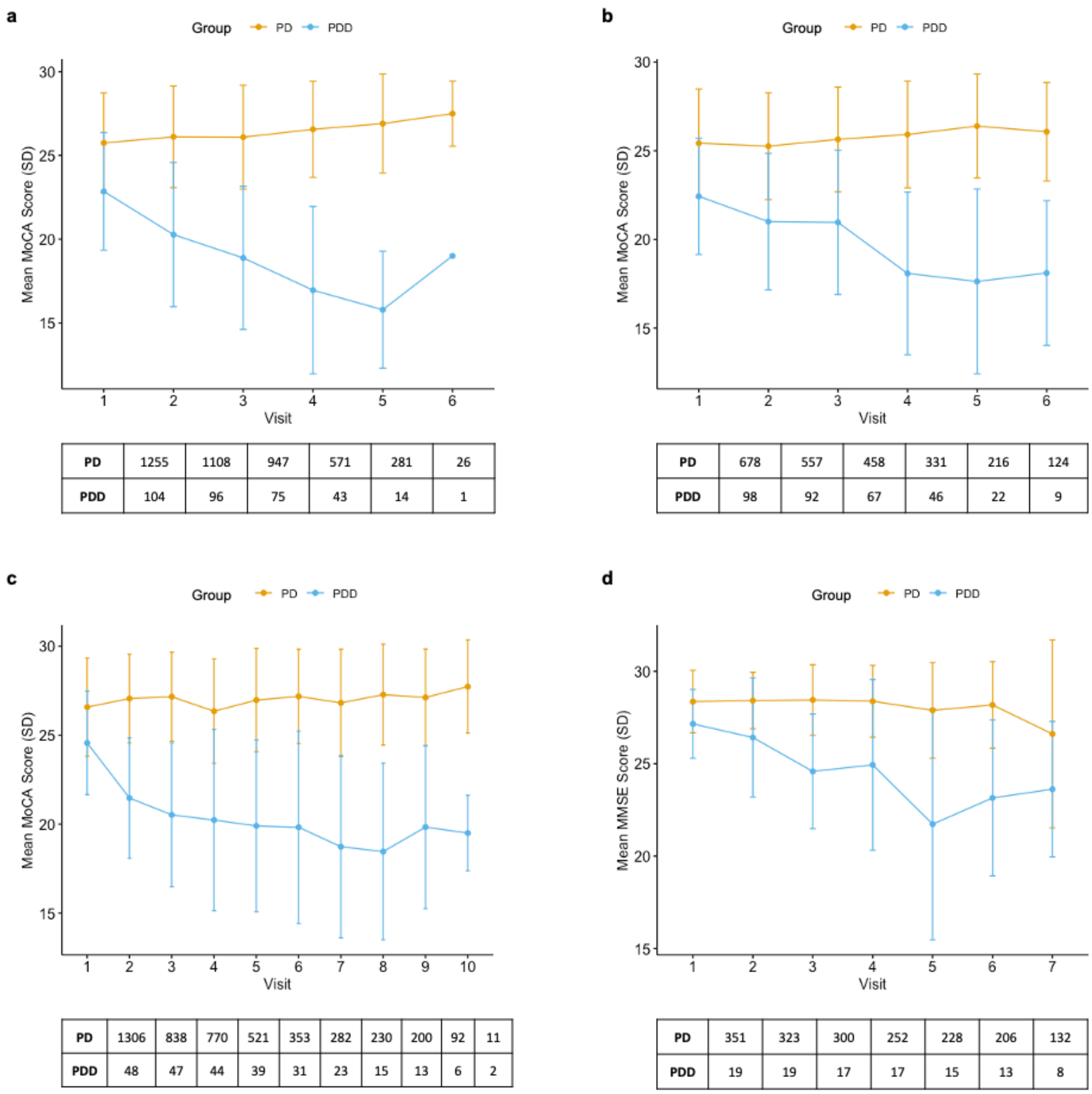


**Supplementary Fig. 2: Cognitive scores and number of assessments over time in PD and PDD cases. a-c**, MoCA scores (top panels) and number of assessments (bottom panels) at each study time point in TPD (**a**), OPDC (**b**) and AMP-PD (**c**) cohorts. **d**, MMSE scores (top panel) and number of assessments (bottom panel) at each study time point in the DIGPD cohort. Mean ± standard deviation.

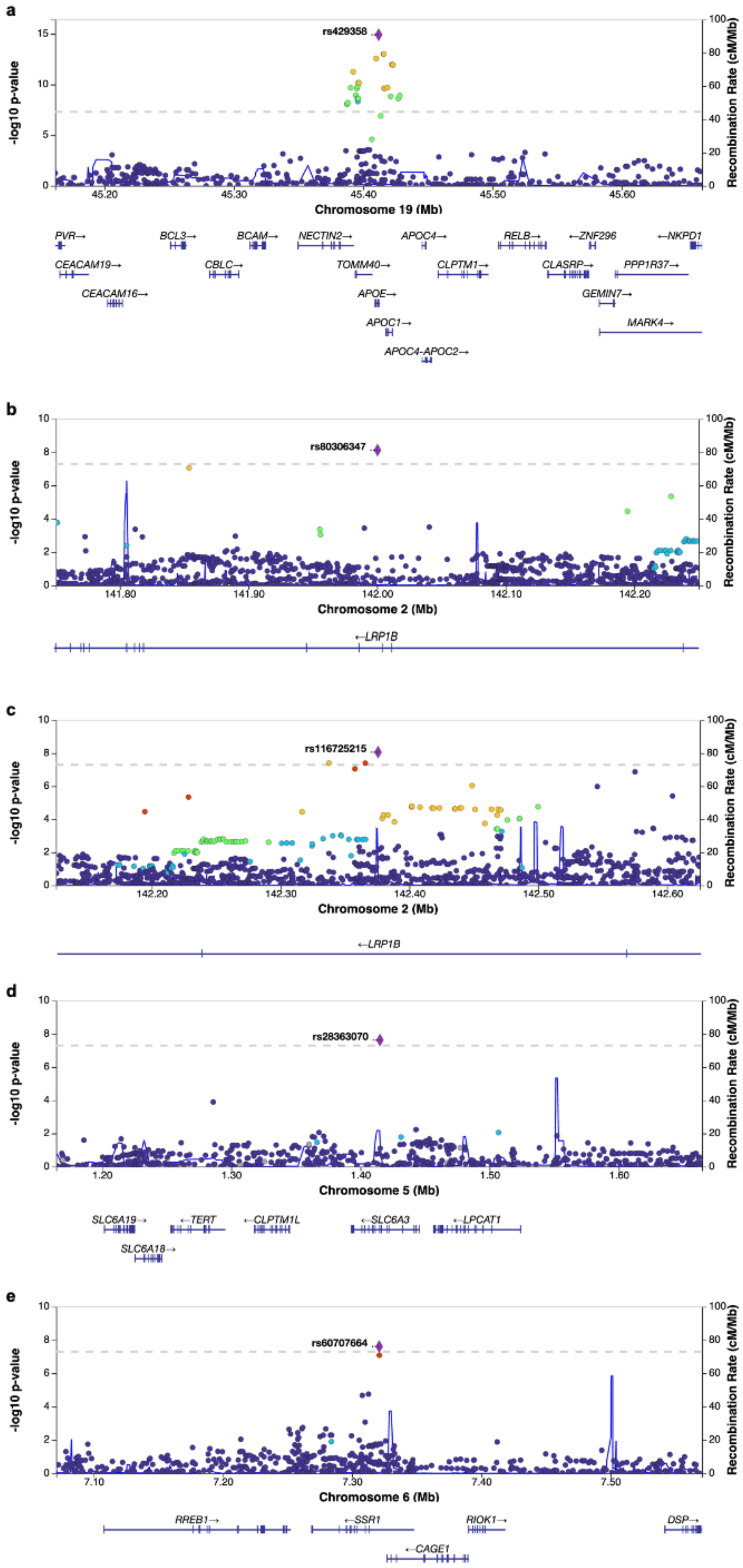
a
rs429358
-log10 p-value
Recombination Rate (cM/Mb)
Chromosome 19 (Mb)
45.20
45.30
45.40
45.50
45.60
PVR→
BCL3→
BCAM→
NECTIN2→
APOC4→
RELB→
←ZNF296
←NKPD1
CEACAM19→
CBLC→
TOMM40→
CLPTM1→
CLASRP→
PPP1R37→
CEACAM16→
APOE→
GEMIN7→
APOC1→
MARK4→
APOC4-APOC2→
b
rs80306347
-log10 p-value
Recombination Rate (cM/Mb)
141.80
141.90
142.00
142.10
142.20
Chromosome 2 (Mb)
←LRP1B
c
rs116725215
-log10 p-value
Recombination Rate (cM/Mb)
142.20
142.30
142.40
142.50
142.60
Chromosome 2 (Mb)
←LRP1B
d
rs28363070
-log10 p-value
Recombination Rate (cM/Mb)
1.20
1.30
1.40
1.50
1.60
Chromosome 5 (Mb)
SLC6A19→
←TERT
←CLPTM1L
←SLC6A3
←LPCAT1
SLC6A18→
e
rs60707664
-log10 p-value
Recombination Rate (cM/Mb)
7.10
7.20
7.30
7.40
7.50
Chromosome 6 (Mb)
RREB1→
←SSR1
RIOK1→
DSP→
←CAGE1

**Supplementary Fig. 3: Regional association plots and recombination rates at the genome-wide significant *loci*. a-e**, Regional association plots at 19:45411941 (rs429358) (**a**), 2:142000271 (rs80306347) (**b**), 2:142375694 (rs116725215) (**c**), 5:1415068 (rs28363070) (**d**), and 6:7320911 (rs60707664) (**e**). The index variants are indicated by a purple diamond and their rsID. A measure of linkage disequilibrium between the index variant and variants in their vicinity ($r^2$) is color-coded (dark blue: $0 \le r^2 < 0.2$; light blue: $0.2 \le r^2 < 0.4$; green: $0.4 \le r^2 < 0.6$; orange: $0.6 \le r^2 < 0.8$; red: $0.8 \le r^2 \le 1$; gray: no $r^2$ available). Plots were generated in http://locuszoom.org/.

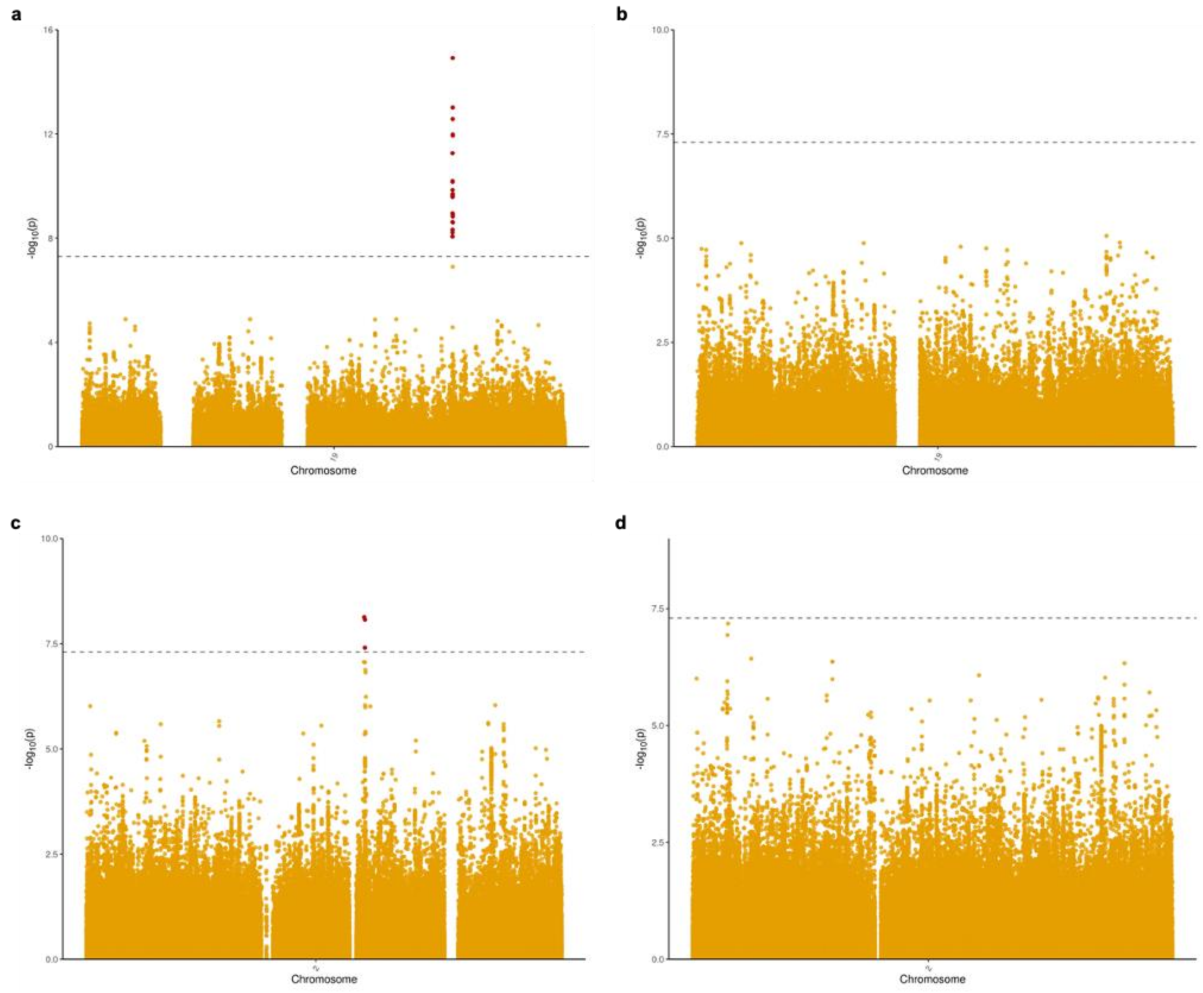


**Supplementary Fig. 4: Conditional analysis of *APOE* and *LRP1B loci*.** **a**, Unconditioned Manhattan plot of chromosome 19. **b**, Manhattan plot of chromosome 19 conditioned on the *APOE* variant rs429358. **c**, Unconditioned Manhattan plot of chromosome 2. **d**, Manhattan plot of chromosome 2 conditioned on the *LRP1B* variant rs80306347. Conditional analysis was performed using the summary statistics of the GWSS meta-analysis in GCTA-COJO while conditioning on a single SNP.

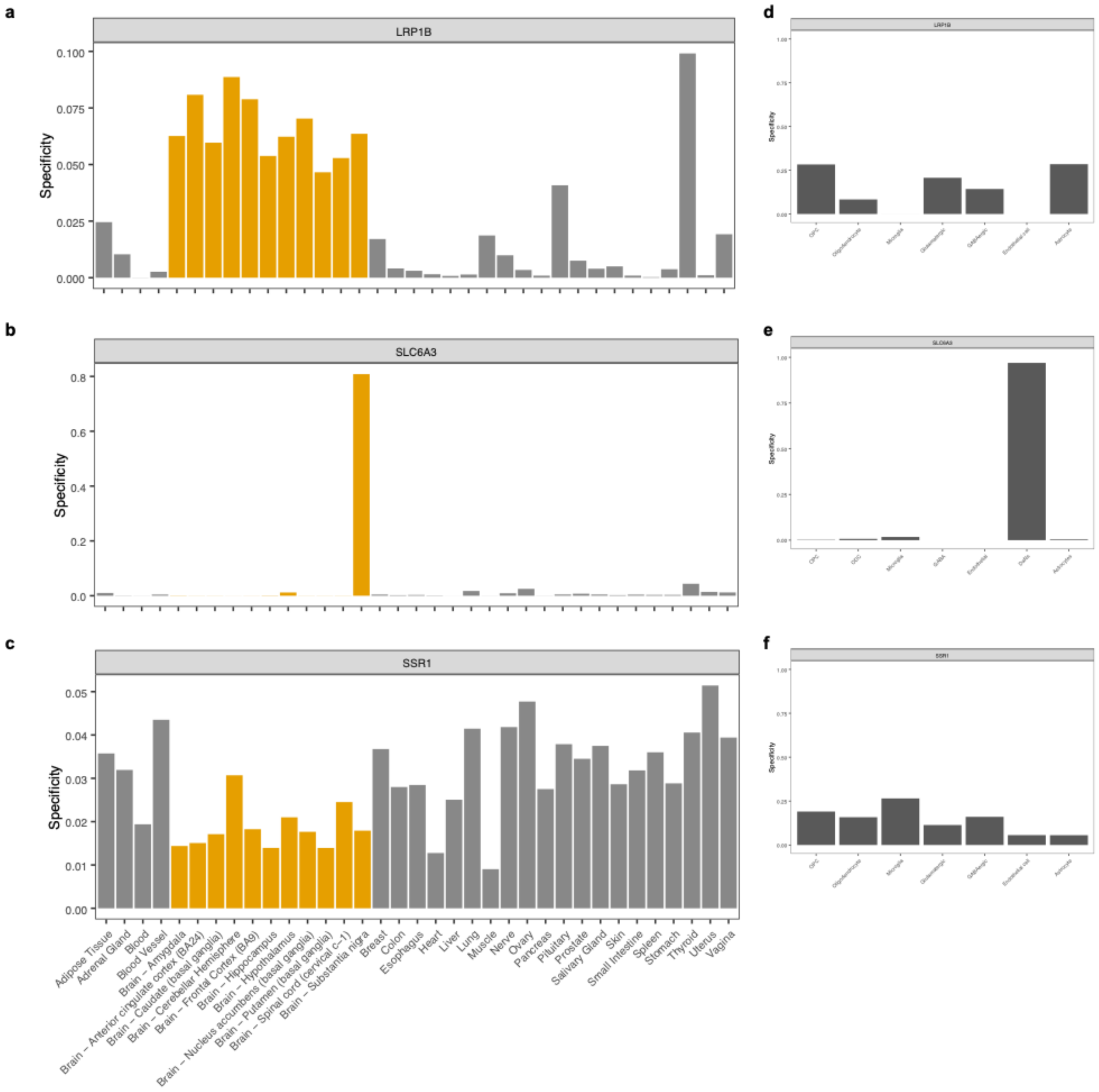


**Supplementary Fig. 5: Tissue and single-cell specificity of novel GWSS hits. a-c**, Plots of *LRP1B* (**a**)*, SLC6A3* (**b**) and *SSR1* (**c**) tissue specificity (GTEx v8 dataset). Bars in orange indicate brain tissues. **d, f**, Plots of *LRP1B* (**d**) and *SSR1* (**f**) cell-type specificity (Allen Brain Atlas single cell gene expression dataset). **e**, Plot of *SLC6A3* cell-type specificity (data from single-cell atlas of human substantia nigra, Agarwal et al., 2020). A specificity value of 0 indicates a gene is not expressed in a cell type/tissue, while a specificity value of 1 indicates that a gene is only expressed in that cell type/tissue. *OPC,* oligodendrocyte precursor cells; *ODC* oligodendrocytes; *GABA,* GABAergic interneurons; *DaNs* dopaminergic neurons.

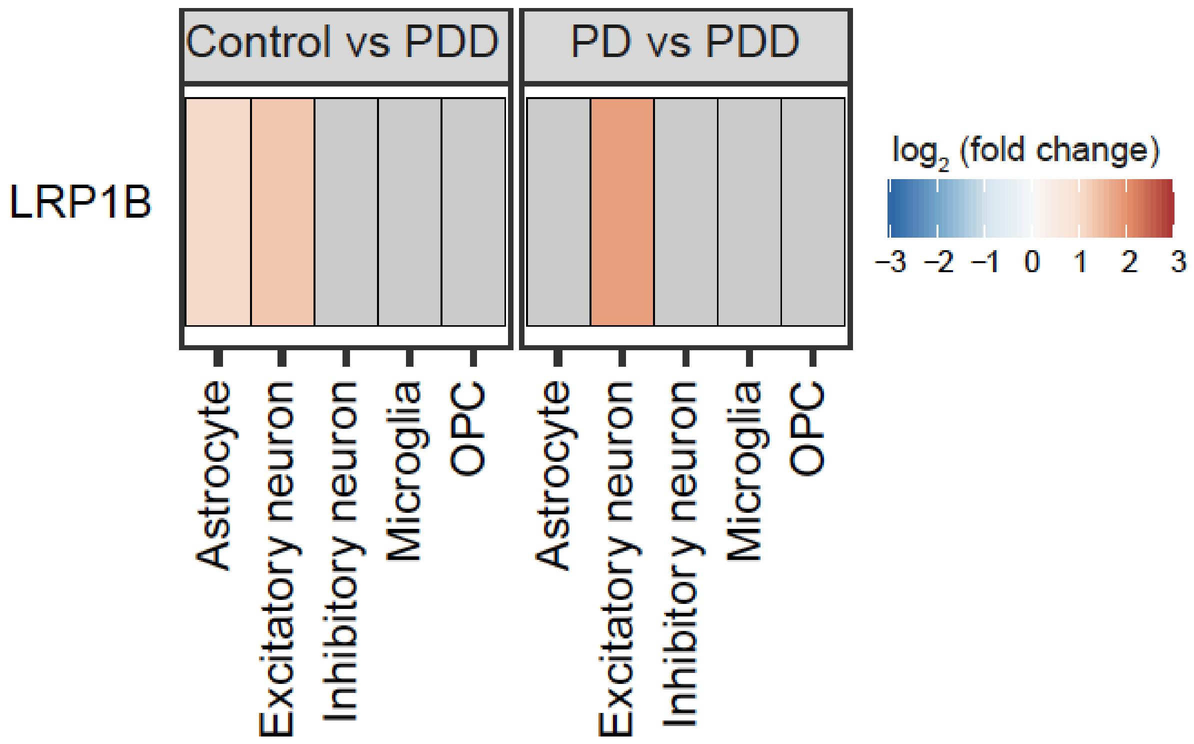


**Supplementary Fig. 6: Differential expression of *LRP1B*.** Representation of differential expression of *LRP1B* across cell types and pairwise comparisons of PDD, PD and the control group based on single-nucleus RNA-sequencing data of the anterior cingulate cortex from Feleke et al. (2021). Genes were considered differentially expressed if FDR < 0.05 and the absolute fold change greater than 1.5. Fill of the tile indicates the $\log_2$(fold change), with non-significant results (FDR > 0.05) coloured grey. *LRP1B* was not differentially expressed in any cell types for the comparison Control vs PD, thus this comparison is not plotted.

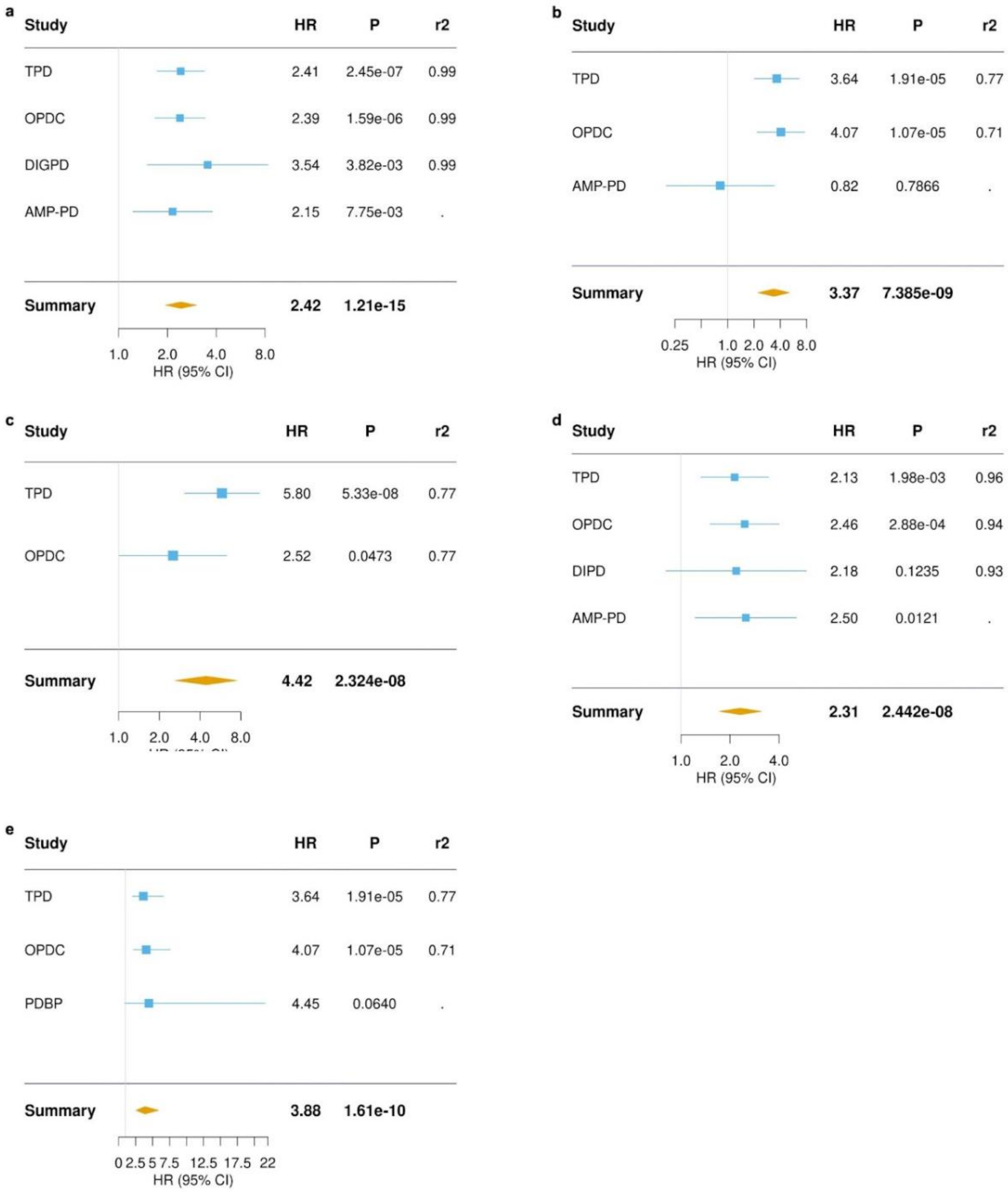


**Supplementary Fig. 7: Forest plots of top hits of GWSS meta-analysis. a**, *APOE* rs429358-C variant ($I^2$ = 0.0; Cochran's Q test: $\chi^2$ = 0.927, df = 3, *P* = 0.8188). **b**, *LRP1B* rs80306347-C variant ($I^2$ = 28.5; $\chi^2$ = 4.195, df = 3, *P* = 0.2412). **c**, *SLC6A3* rs28363070-A variant ($I^2$ = 0.0; $\chi^2$ = 2.155, df = 3, *P* = 0.5408). **d**, rs60707664-A variant near *SSR1* ($I^2$ = 0.0; $\chi^2$ = 0.228, df = 3, *P* = 0.9733). **e**, *LRP1B* rs80306347-C variant after meta-analysis of individual AMP-PD cohorts ($I^2$ = 0.0; $\chi^2$ = 0.094, df = 4, *P* = 0.9989). Results are not available for the DIGPD cohort in **b-c** and **e**, for the AMP-PD cohort in **c,** and for PPMI and SURE-PD3 sub-cohorts in **e** due to no events occurring in variant carriers generating infinite estimates in the Cox proportional hazards analysis. *HR*, hazard ratio; *CI*, confidence interval; *P*, *p*-value; *r2*, imputation info score.

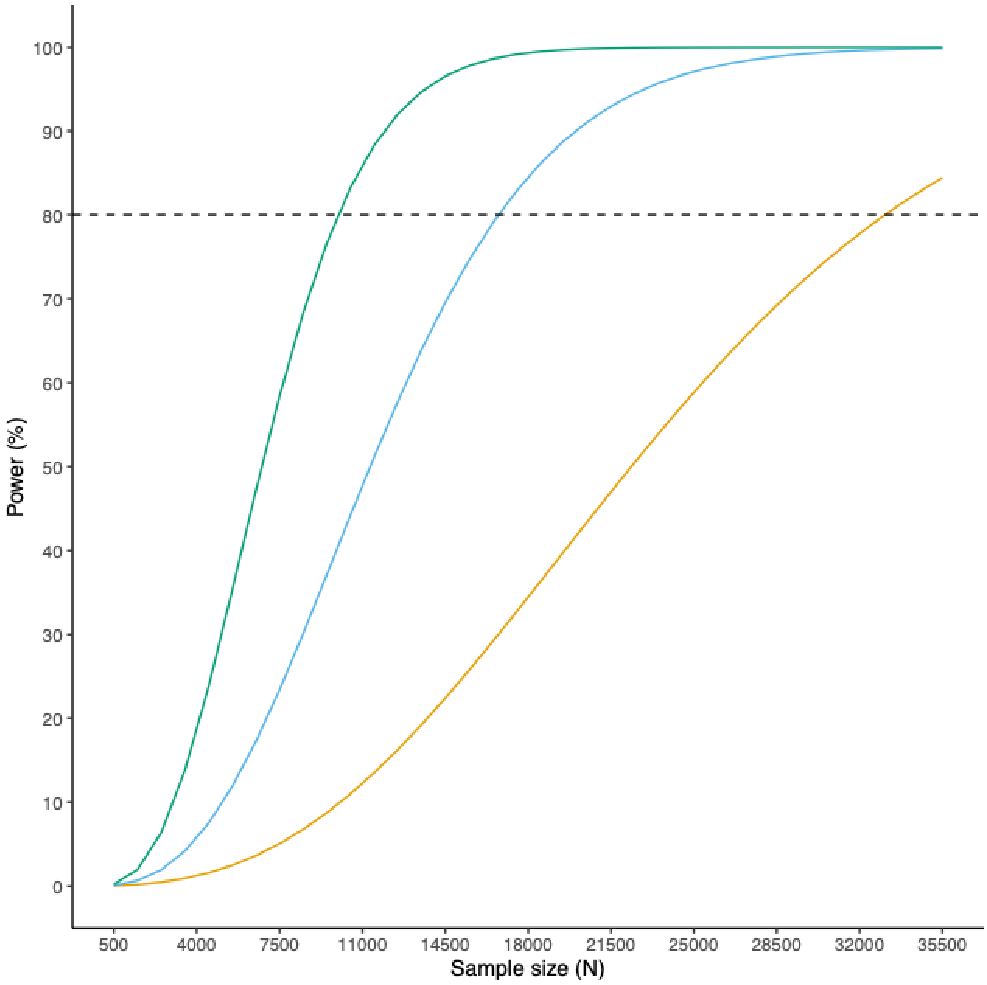


**Supplementary Fig. 8: Plot of statistical power modelling.** Modelling was performed across a range of event rates for a hypothetical SNP with MAF=0.02 and HR=2, considering the mean time-to-event of 4.5 years. The colours represent the event rates of the AMP-PD (orange), TPD (blue) and OPDC (green) cohorts. A dashed line indicates 80% statistical power.

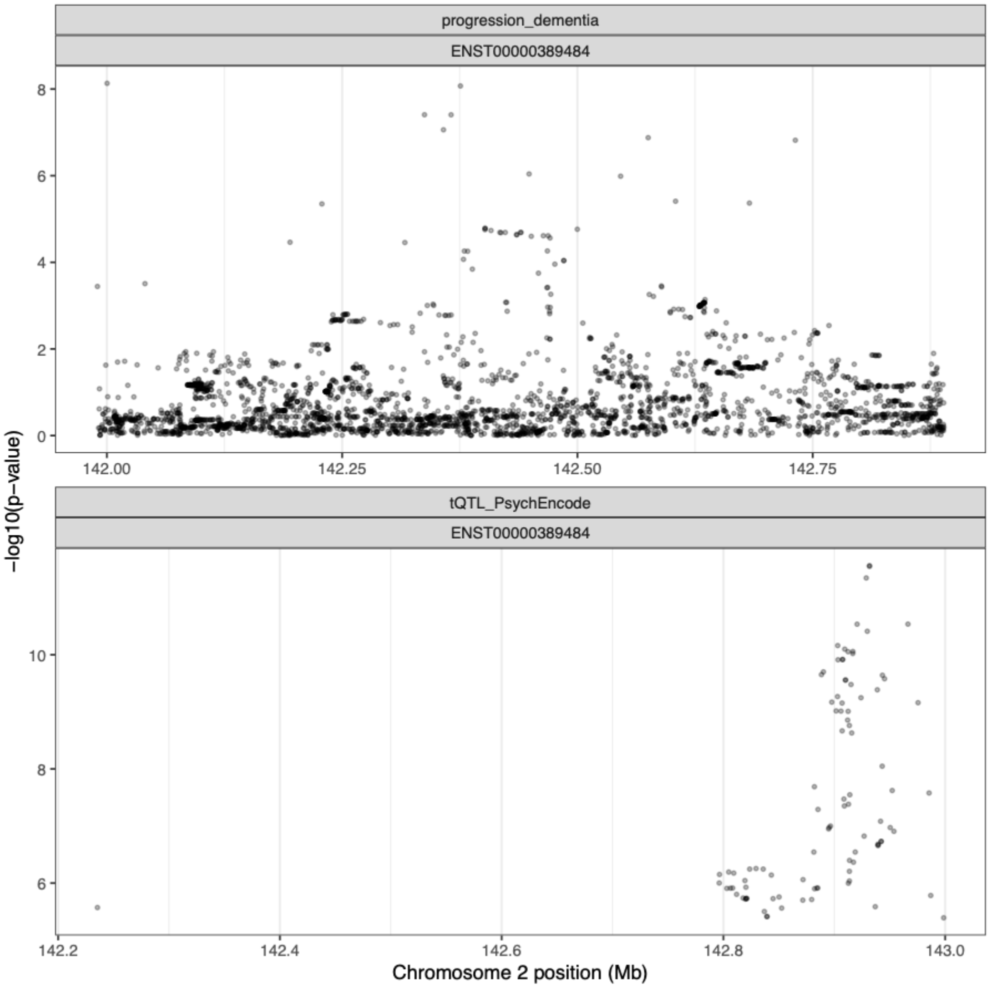


**Supplementary Fig. 9: Regional association plots for tQTL and *LRP1B* GWSS.** Plots were generated using a FDR-filtered tQTL dataset from PsychENCODE.

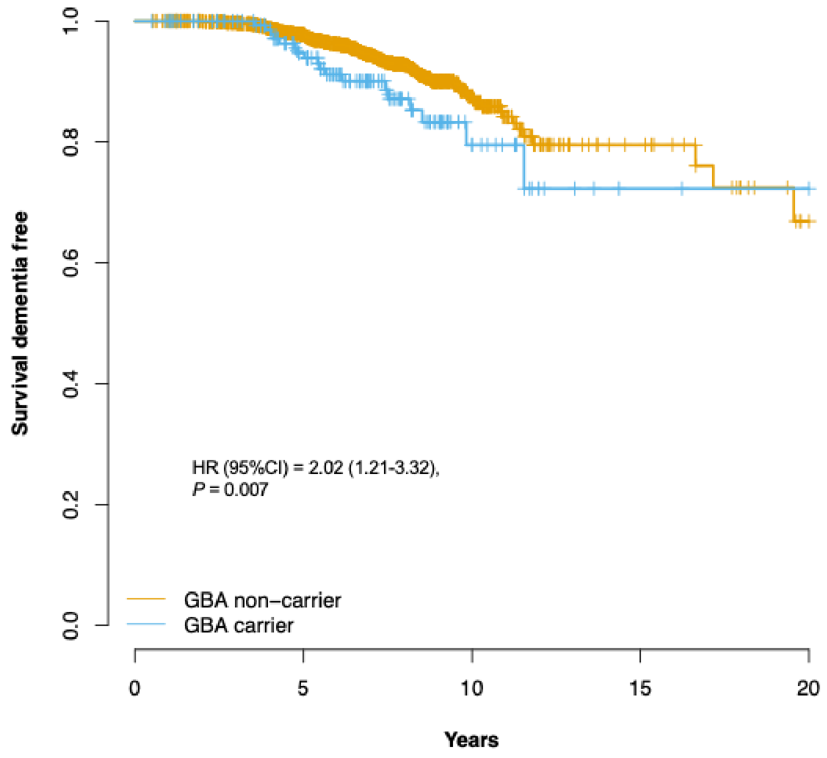


**Supplementary Fig. 10: Survival curves by *GBA* status.** *GBA* mutations determined by Sanger sequencing of subset of samples from DIGPD and TPD ($n$ = 1,793). A Cox proportional hazards model adjusted for gender, age at onset, first 5 PCs and a cohort term was used to analyse the contribution of *GBA* mutations to the rate of progression to PDD. Carriers of GD-causing mutations (D380A, L444P, N409S, R463C, G202R, R359*, G377S, F213I, R257Q, Rec 1 allele (L444P, A456P, V460V), Rec2 allele (D409H, L444P, A456P, V460V), D140H, Rec L444P + A456P) and PD-risk polymorphisms (E365K, T408M) were combined as a single group for analysis. Individuals with variants of uncertain significance were excluded from the analysis. *HR*, hazard ratio; *CI*, confidence interval; *P*, *p*-value.

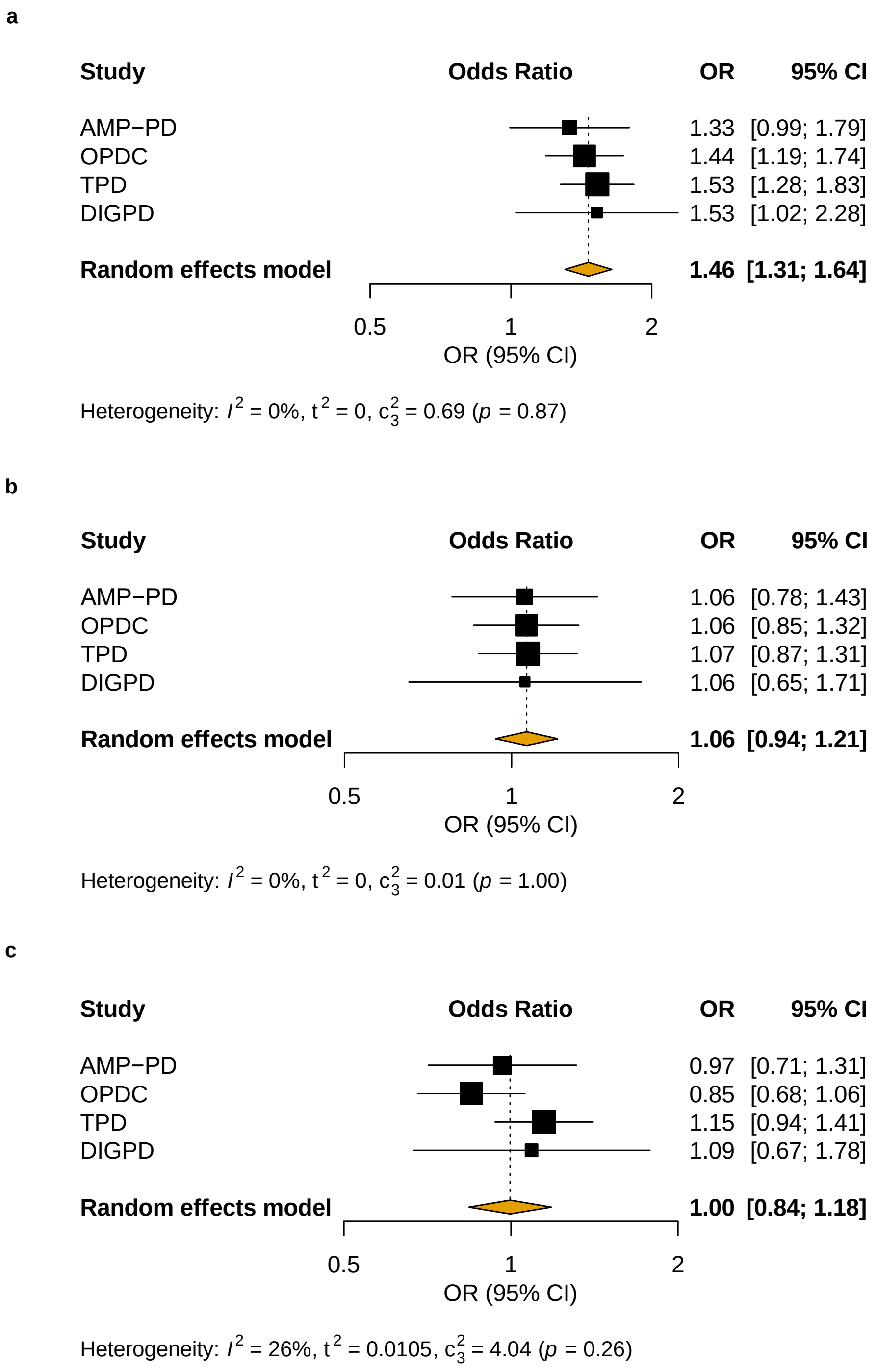


**Supplementary Fig. 11: Forest plots of genetic risk scores meta-analysis. a**, Forest plot of the meta-analysis of the Alzheimer’s disease genetic risk score in PD and PDD cases based on summary statistics of Kunkle et al. (2019). **b**, Forest plot of the meta-analysis of the Alzheimer’s disease genetic risk score in PD and PDD cases after exclusion of the APOE variant 19:45411941 T>C (rs429358) from the input file. **c**, Forest plot of the meta-analysis of the Parkinson’s disease genetic risk score score in PD and PDD cases based on summary statistics of Nalls et al. (2019). *OR*, odds ratio; *CI*, confidence interval.

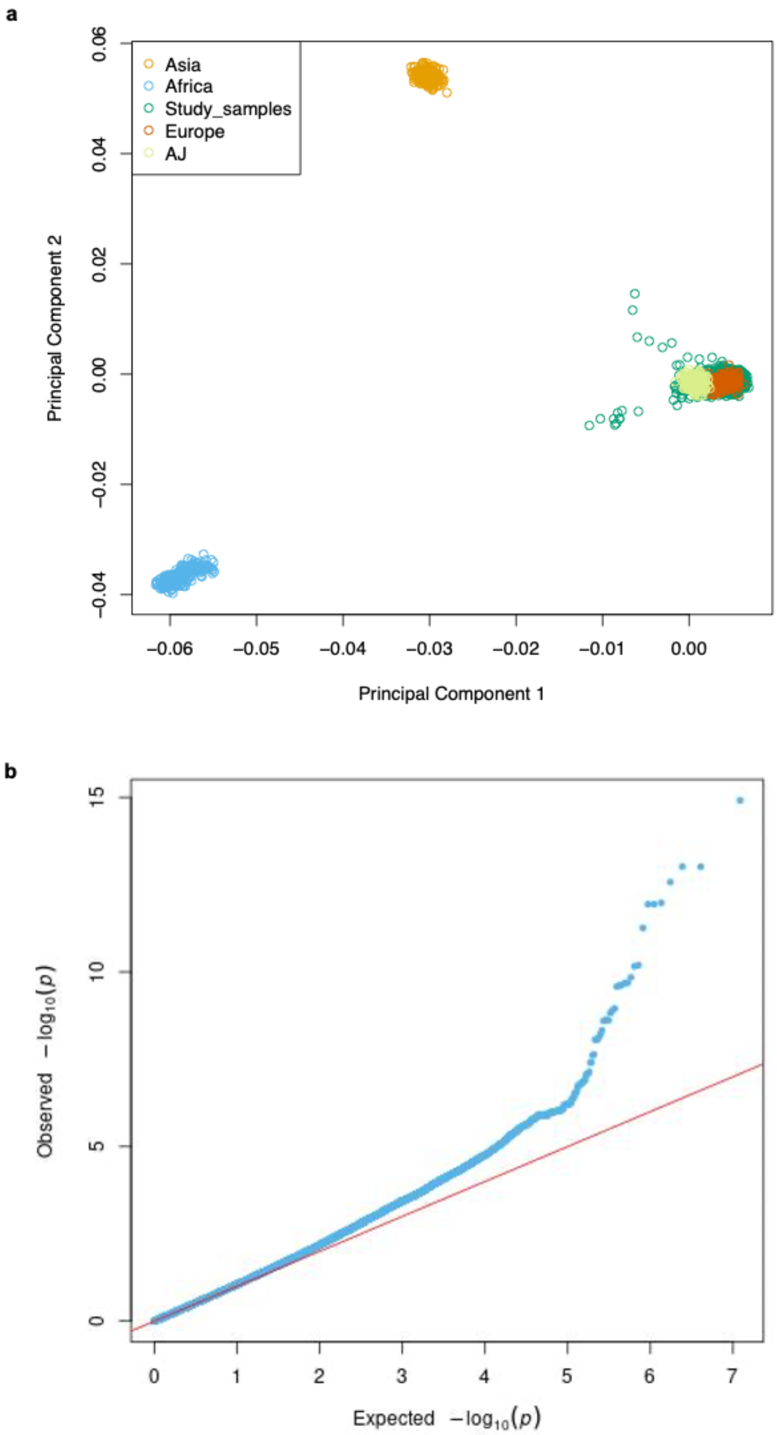


**Supplementary Fig. 12: Principal components and quantile-quantile plots. a**, First two principal components (PCs) of the combined datasets plotted against the HapMap3 Genome Reference Panel and the Ashkenazi Jewish population panel (accessible through the Gene Expression Omnibus (GEO) database [https://www.ncbi.nlm.nih.gov/gds], accession number GSE23636). **b**, Quantile-quantile (QQ) plot of the GWSS meta-analysis. The genomic inflation factor ($\lambda$gc) was 1.048.

### Supplementary Table 1. Demographic characteristics per cohort

| Cohort | N (% female) | AAO* | AAB | Baseline MoCA | Baseline MMSE | Disease duration to baseline (Years) | Education ≤ 12 years (%) | Event rate (%) | Time PD onset to PDD (Years) | Time in study (Months, median) |
|---|---|---|---|---|---|---|---|---|---|---|
| TPD | 1454 (35.4) | 64.0±9.7 | 67.1±9.1 | 25.5±3.1 | n/a | 3.1±3.0 | 31.2 | 7.3 | 6.6±3.37 | 44.2 |
| OPDC | 782 (35.2) | 64.3±9.6 | 67.1±9.5 | 25.1±3.2 | n/a | 2.84±1.82 | 38.9 | 12.7 | 6.8±2.84 | 53.6 |
| DIGPD | 370 (39.7) | 58.7±10 | 62.2±9.9 | n/a | 28.3±1.73 | 3.44±1.56 | 37.8 | 5.1 | 7.7±2.58 | 60.8 |
| AMP-PD^ | 1358 (38.1) | 60.0±9.7 | 63.7±9.1 | 26.5±2.8 | n/a | 3.71±4.71 | 12.4 | 3.6 | 5.8±4.03 | 30 |

Mean ± standard deviation are provided, except where indicated.
*In AMP-PD, only age at diagnosis is available.
^BioFIND study (*n* = 88); PDBP study (*n* = 671); PPMI study (*n* = 368); SURE-PD3 study (*n* = 231).
*AAO,* age at PD onset; *AAB*, age at baseline; *MoCA*, Montreal Cognitive Assessment; *MMSE*, Mini-Mental State Examination; *n/a*, non-applicable.

### Supplementary Table 2. Conditional analysis on top *APOE* signals

| | SNP | Nearest gene | Effect allele | HR | 95% CI | *P*-value |
|---|---|---|---|---|---|---|
| Unconditioned | 19:45411941 | APOE | C | 2.42 | 1.95 - 3.01 | 1.21e-15 |
| | 19:45396219 | TOMM40 | T | 1.93 | 1.58 - 2.35 | 6.39e-11 |
| | 19:45428234 | APOC1P1 | A | 1.94 | 1.57 - 2.40 | 1.24e-09 |
| | 19:45396144 | TOMM40 | T | 1.99 | 1.59 - 2.50 | 2.40e-09 |
| Conditioned on rs429358 (19:45411941 T>C) | 19:45396219 | TOMM40 | T | 1.08 | 0.89 - 1.32 | 0.4257 |
| | 19:45428234 | APOC1P1 | A | 1.02 | 0.82 - 1.27 | 0.8372 |
| | 19:45396144 | TOMM40 | T | 1.03 | 0.82 - 1.29 | 0.7889 |

*HR*, hazard ratio; *CI*, confidence interval.
Genome-wide significance level set at $5x10^{-08}$.

### Supplementary Table 3. Conditional analysis of top *LRP1B* signal

| | SNP | Effect allele | HR | 95% CI | *P-value* |
|---|---|---|---|---|---|
| Unconditioned | 2:142375694 G>T | T | 3.48 | 2.28 - 5.32 | 8.53e-09 |
| | 2:142000271 T>C | C | 3.37 | 2.23 - 5.09 | 7.34e-09 |
| Conditioned on rs80306347 (2:142000271 T>C) | 2:142375694 G>T | T | 1.81 | 1.19 - 2.77 | 0.0062 |
| Conditioned on rs116725215 (2:142375694 G>T) | 2:142000271 T>C | C | 1.79 | 1.19 - 2.71 | 0.0055 |

*HR*, hazard ratio; *CI*, confidence interval.
Genome-wide significance level set at $5x10^{-08}$.

### Supplementary Table 6. Candidate loci analysis

| | | | | | Effect allele frequency | | | | |
|---|---|---|---|---|---|---|---|---|---|
| **Gene** | **SNP ID** | **Effect allele** | **CHR** | **BP** | **PD** | **PDD** | **HR** | **95% CI** | **Nominal *P*-value** |
| *APOE* | rs429358 | C (ε4) | 19 | 45411941 | 0.1322 | 0.2224 | 2.54 | 1.99-3.24 | 8.16e-14 |
| *GBA* | rs2230288 | T | 1 | 155206167 | 0.0226 | 0.0639 | 2.33 | 1.50-3.61 | 1.64e-4 |
| *MAPT* | rs1800547 | A (H1) | 17 | 44051846 | 0.8157 | 0.8088 | 0.59 | 0.31-1.12 | 0.109 |
| *SNCA* | rs356219* | A | 4 | 90637601 | 0.4253 | 0.3906 | 0.99 | 0.76-1.30 | 0.968 |
| *SNCA* | rs7680557* | A | 4 | 90763360 | 0.4807 | 0.4688 | 1.1 | 0.81-1.49 | 0.534 |
| *SNCA* | rs7681440* | G | 4 | 90756550 | 0.4696 | 0.4531 | 1.07 | 0.79-1.45 | 0.649 |
| *SNCA* | rs11931074 | T | 4 | 90639515 | 0.0983 | 0.0882 | 0.98 | 0.71-1.35 | 0.912 |
| *SNCA* | rs7684318 | C | 4 | 90655003 | 0.0922 | 0.079 | 0.94 | 0.67-1.31 | 0.701 |
| *RIMS2* | rs182987047 | T | 8 | 105249272 | 0.0125 | 0.0202 | 1.36 | 0.74-2.50 | 0.328 |
| *WWOX* | rs8050111 | G | 16 | 78281160 | 0.0653 | 0.0643 | 0.94 | 0.66-1.35 | 0.7476 |
| *TMEM108* | rs138073281 | C | 3 | 132985956 | 0.0207 | 0.0202 | 0.89 | 0.43-1.59 | 0.591 |

Cox PH models adjusted for age at onset, sex, first 5 PCs and a cohort term were used to study the association of candidate loci and progression to PDD in the combined cohort (*n* = 3694).
*SNP not available in the AMP-PD cohort (*n* = 2830).
*CHR*, chromosome; *BP,* base pair position in hg19; *HR*, hazard ratio, *CI*, confidence interval.
Significance level was defined as **α**=0.00455 after Bonferroni correction for multiple testing (0.05/11).

### Supplementary Table 7. GBA mutations in DIGPD and TPD cohorts

| GD pathogenic mutations | PD risk variants | Rare variants of unknown significance | N patients (%) |
|---|---|---|---|
| L444P[1] | | | 20 (1.1) |
| N409S | | | 15 (0.8) |
| R463C | | | 5 (0.3) |
| G202R | | | 2 (0.1) |
| R359*[2] | | | 2 (0.1) |
| G377S | | | 2 (0.1) |
| R257Q | | | 1 (0.1) |
| D409H | | | 1 (0.1) |
| Rec1 (L444P, A456P, V460V) | | | 4 (0.2) |
| Rec2 (D409H, L444P, A456P, V460V) | | | 1 (0.1) |
| Rec L444P + A456P | | | 1 (0.1) |
| D179H[3] | | | 2 (0.1) |
| N409S/L444P[4] | | | 2 (0.1) |
| | E365K | | 77 (4.3) |
| | T408M | | 32 (1.8) |
| | | L66P, S173s, G10S, R170H, L175I, P55S, R329H, R395C, M123T, L383Xfs, L105R, Ex4 hemizygous deletion, P266Xfs, T267I/L268L, G189V | 16 (0.9) |

A total of 2,455 cases were Sanger sequenced for GBA, of which 1,793 overlapped with the individuals included in the GWSS. Individuals with variants of unknown significance were removed from the analysis. [1]L444P/E365K (*n* = 2), [2]R359*/T408M (*n* = 1), [3]D179H/E365K (*n* = 2), [4]N409S/L444P/T408M (*n* = 1).

### Supplementary Table 8. Demographic characteristics of left censored individuals per cohort

| Cohort | N | AAO* | AAB | Baseline MoCA | Baseline MMSE | Disease duration to baseline (Years) | Education ≤ 12 years (%) |
|---|---|---|---|---|---|---|---|
| TPD | 35 | 69.3 ± 8.3 | 73.6 ± 7.7 | 16.3 ± 3.1 | n/a | 3.9 ± 3.34 | 38.5 |
| OPDC | 13 | 71.6 ± 8.9 | 74.5 ± 8.0 | 18 ± 1.5 | n/a | 2.91 ± 1.49 | 46.2 |
| DIGPD | 3 | 69.1 ± 8.2 | 74.3 ± 8.2 | n/a | 22.7 ± 1.5 | 5.18 ± 0.34 | 0 |
| AMP-PD | 21 | 59.6 ± 9.7 | 70.9 ± 6.7 | 15.5 ± 4.6 | n/a | 10.95 ± 7.23 | 4.8 |

Mean ± standard deviation are provided, except where indicated.
*In AMP-PD, only age at diagnosis is available.
*AAO,* age at PD onset; *AAB*, age at baseline; *MoCA*, Montreal Cognitive Assessment; *MMSE*, Mini-Mental State Examination; *n/a*, non-applicable.

**DIGPD Study group**

**Steering committee:** Jean-Christophe Corvol, MD, PhD (Pitié-Salpêtrière Hospital, Paris, principal investigator of DIGPD), Alexis Elbaz, MD, PhD (CESP, Villejuif, member of the steering committee), Marie Vidailhet, MD (Pitié-Salpêtrière Hospital, Paris, member of the steering committee), Alexis Brice, MD (Pitié-Salpêtrière Hospital, Paris, member of the steering committee and PI for genetic analysis).

**Statistical analyses:** Alexis Elbaz, MD, PhD (CESP, Villejuif, PI for statistical analyses), Fanny Artaud, PhD (CESP, Villejuif, statistician).

**Principal investigators for sites (alphabetical order):** Frédéric Bourdain, MD (CH Foch, Suresnes, PI for site), Jean-Philippe Brandel, MD (Fondation Rothschild, Paris, PI for site), Jean-Christophe Corvol, MD, PhD (Pitié-Salpêtrière Hospital, Paris, PI for site), Pascal Derkinderen, MD, PhD (CHU Nantes, PI for site), Franck Durif, MD (CHU Clermont-Ferrand, PI for site), Richard Levy, MD, PhD (CHU Saint-Antoine, Paris, PI for site), Fernando Pico, MD (CH Versailles, PI for site), Olivier Rascol, MD (CHU Toulouse, PI for site).

**Co-investigators (alphabetical order):** Anne-Marie Bonnet, MD (Pitié-Salpêtrière Hospital, Paris, site investigator), Cecilia Bonnet, MD, PhD (Pitié-Salpêtrière Hospital, Paris, site investigator), Christine Brefel-Courbon, MD (CHU Toulouse, site investigator), Florence Cormier-Dequaire, MD (Pitié-Salpêtrière Hospital, Paris, site investigator), Bertrand Degos, MD, PhD (Pitié-Salpêtrière Hospital, site investigator), Bérangère Debilly, MD (CHU Clermont-Ferrand, site investigator), Alexis Elbaz, MD, PhD (Pitié-Salpêtrière Hospital, Paris, site investigator), Monique Galitsky (CHU de Toulouse, site investigator), David Grabli, MD, PhD (Pitié-Salpêtrière Hospital, Paris, site investigator), Andreas Hartmann, MD, PhD (Pitié-Salpêtrière Hospital, Paris, site investigator), Stephan Klebe, MD (Pitié-Salpêtrière Hospital, Paris, site investigator), Julia Kraemmer, MD (Pitié-Salpêtrière Hospital, site investigator), Lucette Lacomblez, MD (Pitié-Salpêtrière Hospital, Paris, site investigator), Sara Leder, MD (Pitié-Salpêtrière Hospital, Paris, site investigator), Graziella Mangone, MD, PhD (Pitié-Salpêtrière Hospital, Paris, site investigator), Louise-Laure Mariani, MD (Pitié-Salpêtrière

Hospital, Paris, site investigator), Ana-Raquel Marques, MD (CHU Clermont Ferrand, site investigator), Valérie Mesnage, MD (CHU Saint Antoine, Paris, site investigator), Julia Muellner, MD (Pitié-Salpêtrière Hospital, Paris, site investigator), Fabienne Ory-Magne, MD (CHU Toulouse, site investigator), Violaine Planté-Bordeneuve, MD (Henri Mondor Hospital, Créteil, site investigator), Emmanuel Roze, MD, PhD (Pitié-Salpêtrière Hospital, Paris, site investigator), Melissa Tir, MD (CH Versailles, site investigator), Marie Vidailhet, MD (Pitié-Salpêtrière Hospital, Paris, site investigator), Hana You, MD (Pitié-Salpêtrière Hospital, Paris, site investigator).

**Neuropsychologists:** Eve Benchetrit, MS (Pitié-Salpêtrière Hospital, Paris, neuropsychologist), Julie Socha, MS (Pitié-Salpêtrière Hospital, Paris, neuropsychologist), Fanny Pineau, MS (Pitié-Salpêtrière Hospital, Paris, neuropsychologist), Tiphaine Vidal, MS (CHU Clermont-Ferrand, neuropsychologist), Elsa Pomies (CHU de Toulouse, neuropsychologist), Virginie Bayet (CHU de Toulouse, neuropsychologist).

**Genetic core:** Alexis Brice (Pitié-Salpêtrière Hospital, Paris, PI for genetic studies), Suzanne Lesage, PhD (INSERM, ICM, Paris, genetic analyses), Khadija Tahiri, PhD (INSERM, ICM, Paris, lab technician) Hélène Bertrand, MS (INSERM, ICM, Paris, lab technician), Graziella Mangone, MD, PhD (Pitié-Salpêtrière Hospital, Paris, genetic analyses).

**Sponsor activities and clinical research assistants:** Alain Mallet, PhD (Pitié-Salpêtrière Hospital, Paris, sponsor representative), Coralie Villeret (Hôpital Saint Louis, Paris, Project manager), Merry Mazmanian (Pitié-Salpêtrière Hospital, Paris, project manager), Hakima Manseur (Pitié-Salpêtrière Hospital, Paris, clinical research assistant), Mostafa Hajji (Pitié-Salpêtrière Hospital, Paris, data manager), Benjamin Le Toullec, MS (Pitié-Salpêtrière Hospital, Paris, clinical research assistant), Vanessa Brochard, PhD (Pitié-Salpêtrière Hospital, Paris, project manager), Monica Roy, MS (CHU de Nantes, clinical research assistant), Isabelle Rieu, PhD (CHU Clermont-Ferrand, clinical research assistant), Stéphane Bernard (CHU Clermont-Ferrand, clinical research assistant), Antoine Faurie-Grepon (CHU de Toulouse, clinical research assistant).
